# When external validation isn’t enough: Simpson’s paradox, direction asymmetry, and calibration collapse in cross-continental perioperative mortality prediction

**DOI:** 10.64898/2025.12.28.25343118

**Authors:** Debaraj Shome Purkayastha

## Abstract

**Objective:** To test whether stratified within-cohort analysis, bidirectional external validation, and case-level paired bootstrap inference jointly surface failure-mode magnitudes in cross-continental clinical prediction that aggregate metrics conceal.

**Materials and Methods:** Eight machine learning models (XGBoost and logistic regression, on preoperative and preoperative+intraoperative feature sets) were trained on each of INSPIRE (Korea; *n* = 127,413) and MOVER (USA; *n* = 57,545), then evaluated bidirectionally between cohorts. Case-level paired bootstrap (2,000 iterations) was the primary inferential framework. Direction asymmetry was stress-tested via matched-subsampling across four case-mix dimensions (ASA, Elixhauser comorbidity, emergency proportion, temporal period). Feature-importance transferability used SHAP rank correlation; calibration used slope, intercept, O:E, and Brier score before and after Platt scaling.

**Results:** Across the eight cross-population runs, aggregate AUCs concealed substantially lower within-stratum AUCs (Simpson’s paradox gap range 5.0–16.5 pp; worst case: aggregate AUC 0.756 vs within-stratum AUCs 0.58–0.60). Cross-continental transferability was markedly direction-asymmetric (+8.53 pp, 95% CI 6.91–10.24, bootstrap *p* = 0.001); the asymmetry survived matching on ASA and comorbidity, attenuated to 70–86% of baseline after matching emergency proportion, and was untestable for temporal period. Pre-Platt calibration slopes ranged 0.41–1.29 across all eight cross-population runs; 5-fold CV Platt scaling restored slopes to 0.95–1.02. Intraoperative features conferred a mean external AUC advantage of +3.60 pp (95% CI: +2.75 to +4.39 pp; bootstrap p=0.001).

**Discussion:** These magnitudes are clinically material and not visible to conventional aggregate reporting. The methodological commitments that surface them are well-established individually; their joint application here characterizes failure-mode magnitudes that single commitments would underestimate.

**Conclusion:** We present this case study as a cautionary reference for cross-population deployment of clinical prediction models, with reproducibility infrastructure released for verification and extension.

## 1 Background and Significance

Clinical prediction models for perioperative mortality have multiplied rapidly, with internal validation typically reporting strong discrimination (AUC 0.80–0.95) and a 5–10% gain from adding intraoperative vital signs to preoperative risk factors [11, 30]. These models are increasingly being developed and deployed beyond the populations on which they were trained, including in low- and middle-income tertiary surgical centers where deployment-monitoring capacity is most limited [8, 10]. Whether and how their performance survives such transfer has received less empirical attention than internal validation has, even as the TRIPOD+AI statement [6] formalizes contemporary reporting expectations. INSPIRE [15] and MOVER [23] are public perioperative datasets from two academic medical centers on separate continents (Seoul National University Hospital and UC Irvine Medical Center), providing a testbed for population-level transferability with markedly different case-mix profiles (INSPIRE 90% ASA 1–2; MOVER 64% ASA ≥ 3); ASA accordingly provides a clinically meaningful within-stratum stratification variable.

Several aspects of how such models are evaluated have received limited attention. Aggregate discrimination metrics including AUC can overstate clinical utility when models separate patients into risk strata without discriminating within them — an instance of Simpson’s paradox [27], used here in its generalized sense of aggregate metrics misrepresenting within-stratum performance. External validation is typically unidirectional, with directions rarely matched or contrasted, leaving open whether transferability depends on training-population characteristics. Cross-population evidence on which feature classes transfer (and which do not) is scarce, and the algorithmic conditioning of feature-class transfer has not been examined systematically. We report four findings that address these gaps, detailed in §2.3–§2.7.

Aggregate AUC of 0.756 (95% CI: 0.741–0.771) masks within-stratum AUCs near 0.59 for the worst-affected preoperative model, a Simpson’s paradox gap of +16.5 pp (95% CI: +15.1–+18.0 pp). External validation in one direction does not generalize to the other: the unmatched direction-asymmetric gap is +8.53 pp (95% CI: +6.91–+10.24 pp; bootstrap p=0.001) and persists at roughly 70% of that value after multi-dimensional case-mix matching. Two further findings address the underlying mechanism. Intraoperative vital-sign features confer transfer advantage conditional on algorithmic capacity, and SHAP feature-importance ranks transfer near-perfectly across continents while probability calibration does not.

Each of these methodological commitments is well-established individually: stratified within-stratum performance assessment as a response to metric-paradox phenomena in clinical-prediction reporting [5]; case-level resampling for paired comparisons; and external validation as a TRIPOD-checklist item since 2015, with adherence remaining low for prediction models in general [1, 13] and in perioperative medicine specifically [2]. Bidirectional cross-cohort validation is the rarer commitment, requiring paired datasets that remain uncommon. Applied jointly to a cross-continental perioperative testbed, these commitments surface failure-mode magnitudes that single commitments alone would miss.

## 2 Results

### 2.1 Study populations

Two cohorts comprised 127,413 surgical encounters from INSPIRE (Korea, 2011–2020) and 57,545 from MOVER (USA, 2015–2022) after inclusion/exclusion (Figure 1, Table 1). In-hospital mortality was 1,387 (1.1%) (INSPIRE) and 823 (1.4%) (MOVER). The cohorts exhibited substantial case-mix asymmetry (INSPIRE: 90.4% ASA 1–2; MOVER: 63.5% ASA ≥ 3; *p <* 0.001); mortality distribution differed markedly between INSPIRE (deaths spread across the acuity spectrum: 47.2% in ASA 1–2, 52.8% in ASA ≥ 3) and MOVER (concentrated in high-acuity patients: 98.9% in ASA ≥ 3), providing a useful contrast for testing whether training-population characteristics influence transferability.

**Table 1:**
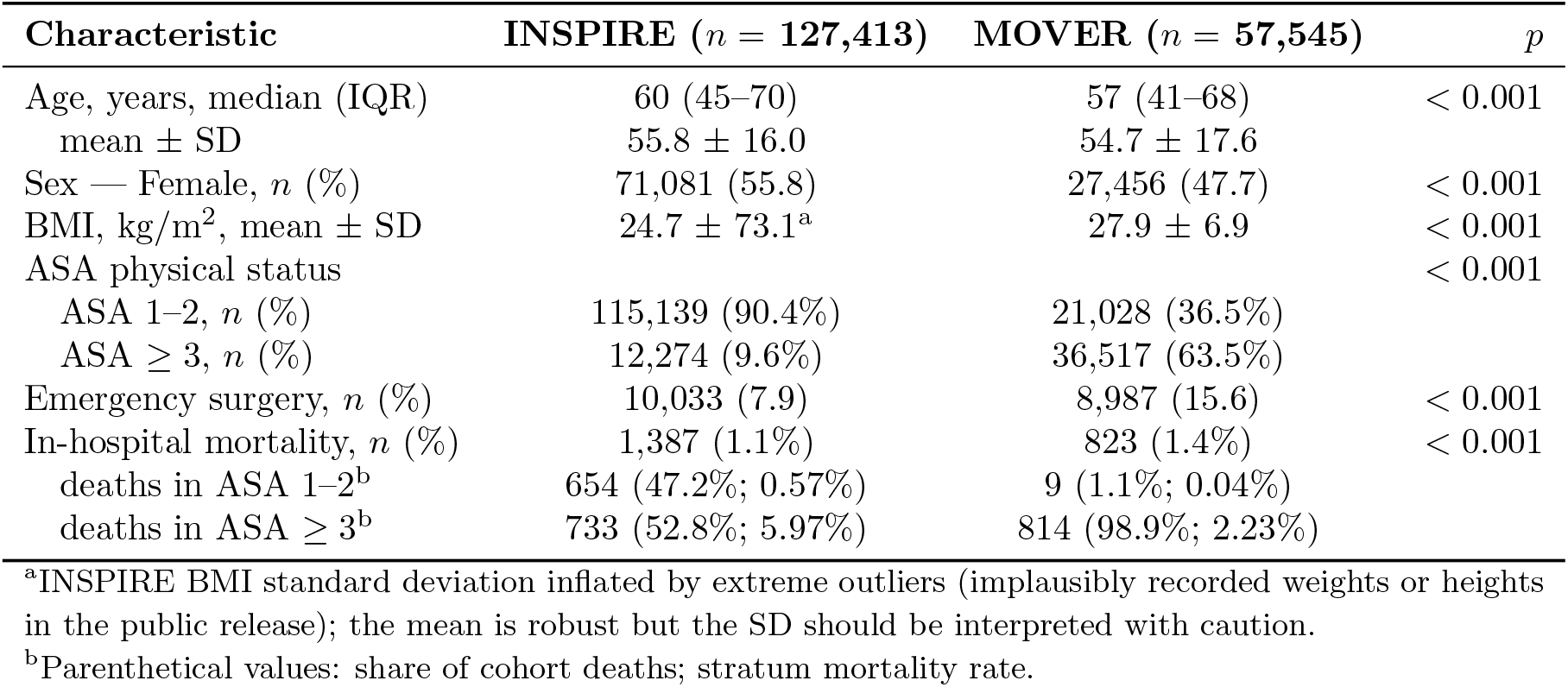
Cohort characteristics. Continuous variables are reported as median (IQR) and mean ± SD; categorical variables as *n* (%). Between-cohort comparisons used Welch’s *t*-test for continuous and *χ*^2^ for categorical variables. The INSPIRE BMI standard deviation (73.1) is outlier-influenced; the median (IQR) form provides a more stable summary.

| Characteristic | INSPIRE ($n = 127,413$) | MOVER ($n = 57,545$) | $p$ |
| --- | --- | --- | --- |
| Age, years, median (IQR) | 60 (45–70) | 57 (41–68) | $< 0.001$ |
| mean ± SD | $55.8 \pm 16.0$ | $54.7 \pm 17.6$ | |
| Sex — Female, $n$ (%) | 71,081 (55.8) | 27,456 (47.7) | $< 0.001$ |
| BMI, kg/m<sup>2</sup>, mean ± SD | $24.7 \pm 73.1^a$ | $27.9 \pm 6.9$ | $< 0.001$ |
| ASA physical status | | | $< 0.001$ |
| ASA 1–2, $n$ (%) | 115,139 (90.4%) | 21,028 (36.5%) | |
| ASA $\geq 3$, $n$ (%) | 12,274 (9.6%) | 36,517 (63.5%) | |
| Emergency surgery, $n$ (%) | 10,033 (7.9) | 8,987 (15.6) | $< 0.001$ |
| In-hospital mortality, $n$ (%) | 1,387 (1.1%) | 823 (1.4%) | $< 0.001$ |
| deaths in ASA 1–2<sup>b</sup> | 654 (47.2%; 0.57%) | 9 (1.1%; 0.04%) |  |
| deaths in ASA $\geq 3$<sup>b</sup> | 733 (52.8%; 5.97%) | 814 (98.9%; 2.23%) | |

<sup>a</sup>INSPIRE BMI standard deviation inflated by extreme outliers (implausibly recorded weights or heights in the public release); the mean is robust but the SD should be interpreted with caution.
<sup>b</sup>Parenthetical values: share of cohort deaths; stratum mortality rate.

**Figure 1:**
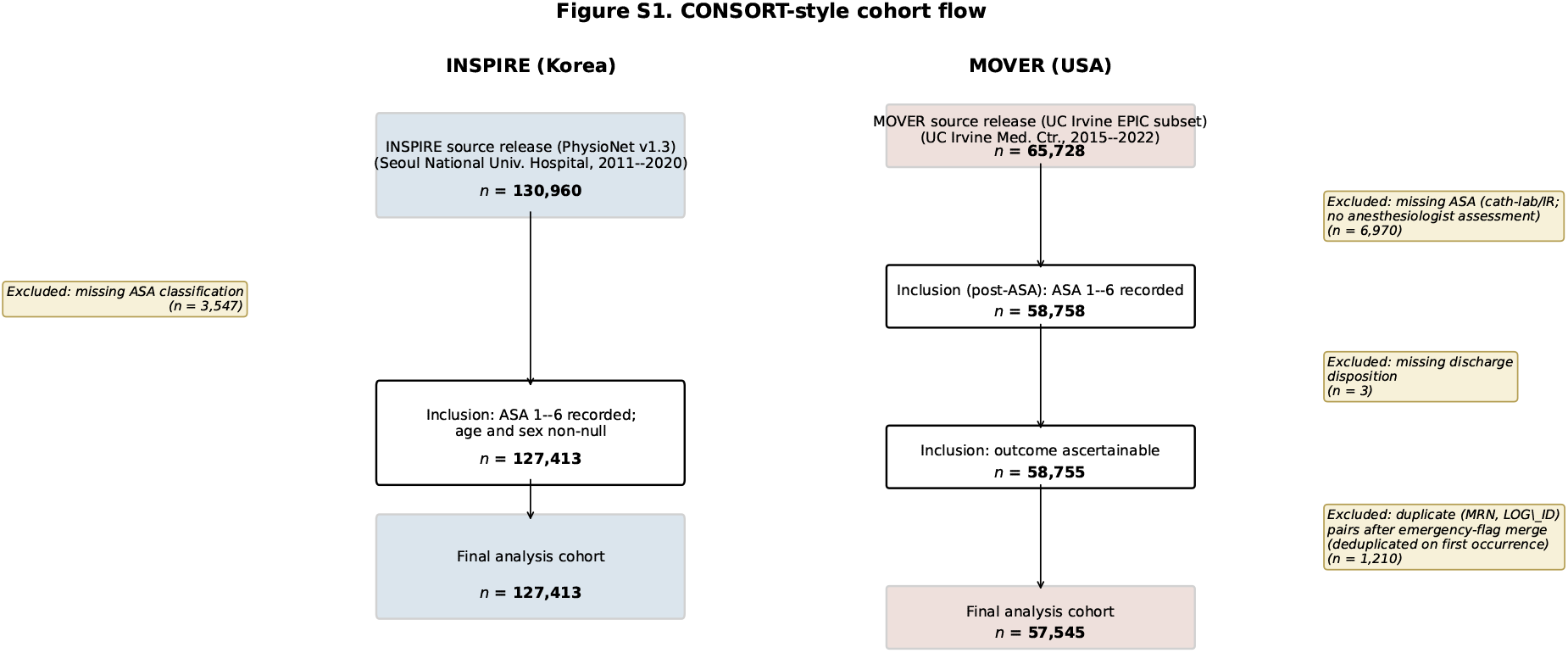
CONSORT-style cohort flow for both perioperative datasets. INSPIRE attrition has a single explicit exclusion step (records missing ASA classification). MOVER attrition has three: an ASA-related exclusion (cardiac-catheterization-laboratory and interventional-radiology procedures with no anesthesiologist ASA assignment), a discharge-disposition exclusion (missing outcome ascertainment), and an (MRN, LOG_ID) deduplication step addressing duplicates introduced by an upstream emergency-flag left-merge. All counts are stated consistently with the cohort summaries reported in Supplementary §S1.1 and §S1.2. *Alt text: CONSORT-style two-pane flow diagram showing how raw records were filtered into the analysis cohorts for INSPIRE (left pane) and MOVER (right pane). INSPIRE shows a single explicit exclusion: 130,876 raw records minus ASA-missing rows yields the 127,413-encounter analysis cohort. MOVER shows three sequential exclusion steps: removal of cardiac-catheterization-laboratory and interventional-radiology procedures lacking anesthesiologist ASA assignment, removal of records with missing discharge disposition, and a deduplication step on the (MRN, LOG ID) key — yielding the 57,545-encounter analysis cohort. Each box shows record count and the exclusion criterion. The diagram makes visible that MOVER attrition is structurally more complex than INSPIRE’s, but that final sample sizes are large*.

### 2.2 Internal validation

All eight models achieved strong internal discrimination under 10-fold stratified cross-validation (out-of-fold concatenated AUC; Supplementary Table S2): INSPIRE-trained models 0.807–0.887, MOVER-trained models 0.914–0.948 (reflecting the stronger discriminative contribution of ASA in a concentrated high-acuity cohort). These internal AUCs serve as the reference point for external-validation degradation throughout.

### 2.3 Simpson’s paradox: aggregate AUC conceals within-stratum discrimination failure

Stratified external validation revealed that aggregate AUC substantially overestimated within-stratum discrimination in preoperative models (Table 2, Figure 2). The most striking example was XGB-MOV-A (XGBoost trained on MOVER with preoperative features only) applied to INSPIRE: overall external AUC was 0.756 (95% CI: 0.741–0.770), yet within-stratum AUC fell to near-random levels (ASA 1–2: 0.597; ASA ≥ 3: 0.584). The resulting paradox gap — defined as overall AUC minus mean within-stratum AUC — was +16.5 pp (95% CI: +15.1–+18.0 pp), the largest observed across the eight models.

**Table 2:** Per-model Simpson’s paradox gap. Columns: training cohort; feature set; overall external AUC; within-stratum AUC mean (or single-stratum fallback for rows with insufficient ASA 1–2 events, marked ^a^); paradox gap (overall − within-stratum) with 95% bootstrap CI. The four ^a^-marked rows had only 9 events in MOVER’s ASA 1–2 stratum, below the *n*_events_ ≥ 10 threshold for stratum-level AUC reliability; their gap is computed as overall AUC − ASA ≥ 3 AUC (single-stratum fallback).

| Model | Trained on | Features | Overall AUC | Within-stratum AUC | Gap (pp) | 95% CI |
| --- | --- | --- | --- | --- | --- | --- |
| XGB-INS-A<sup>a</sup> | INSPIRE | preop | 0.785 | 0.682 | 10.3 | 9.6–10.9 |
| XGB-INS-B<sup>a</sup> | INSPIRE | full (140) | 0.895 | 0.845 | 5.0 | 4.4–5.6 |
| LR-INS-A<sup>a</sup> | INSPIRE | preop | 0.796 | 0.715 | 8.1 | 7.5–8.8 |
| LR-INS-B<sup>a</sup> | INSPIRE | full (140) | 0.741 | 0.687 | 5.4 | 4.9–5.8 |
| XGB-MOV-A | MOVER | preop | 0.756 | 0.590 | **16.5** | 15.1–18.0 |
| XGB-MOV-B | MOVER | full (140) | 0.812 | 0.723 | 8.9 | 7.9–10.0 |
| LR-MOV-A | MOVER | preop | 0.806 | 0.684 | 12.2 | 10.7–13.6 |
| LR-MOV-B | MOVER | full (140) | 0.839 | 0.759 | 8.0 | 6.8–9.2 |

<sup>a</sup>Single-stratum fallback (MOVER ASA 1–2 stratum has $n_{\text{events}} = 9$ ).

**Figure 2:**
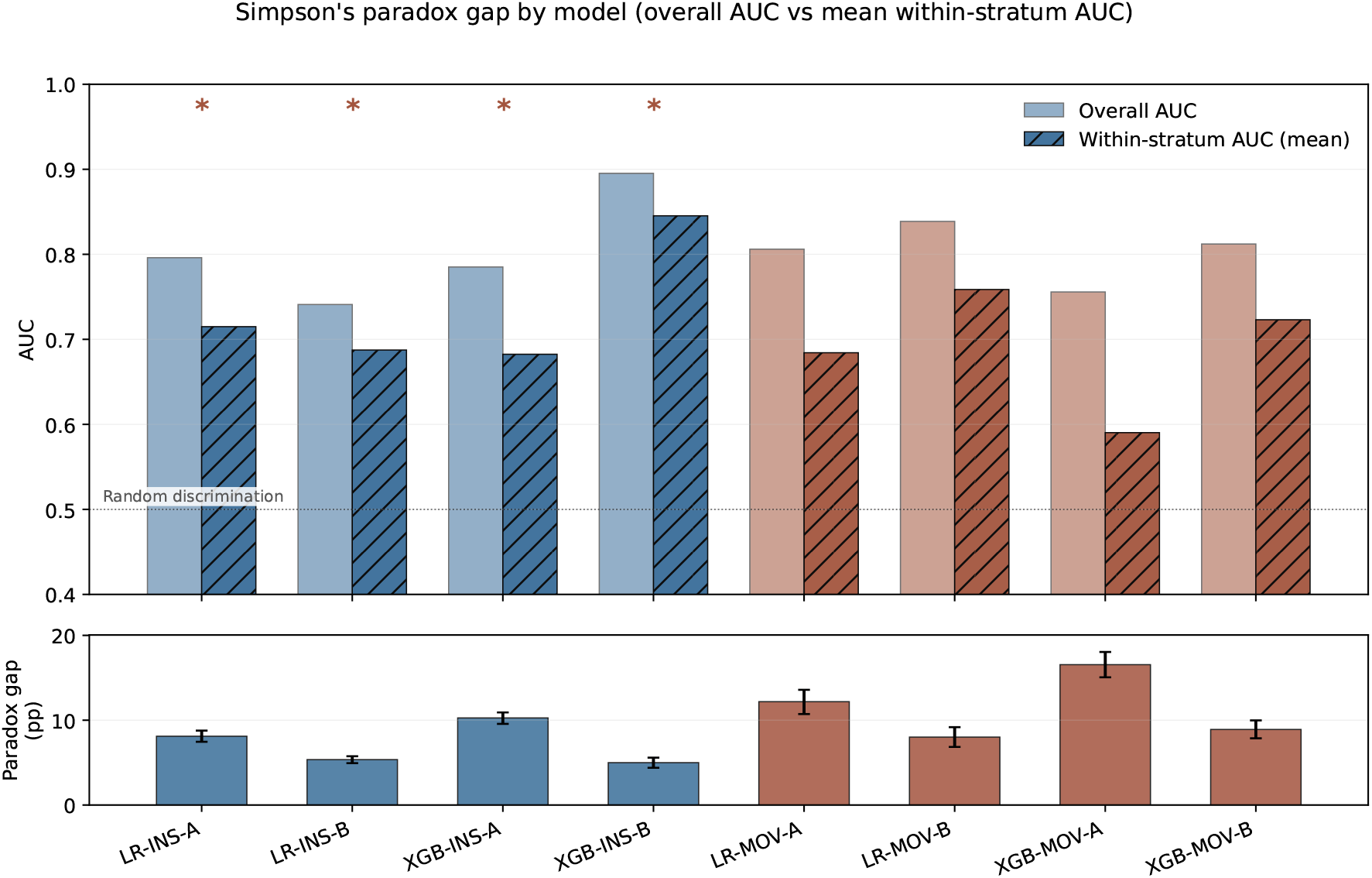
Simpson’s paradox gap across the eight external validations. Top panel: per-model overall AUC (low-saturation) vs. mean within-stratum AUC (hatched). Asterisk markers (**\***) flag the four INSPIRE-trained models on MOVER whose ASA 1–2 stratum has *n*_events_ = 9; for those rows the within-stratum AUC is the ASA ≥ 3 stratum AUC (single-stratum fallback, see Table 2). Bottom panel: paradox gap (overall − within-stratum, pp) with 95% bootstrap CIs. Color: blue = INSPIRE-trained, brown = MOVER-trained. The largest gap is XGB-MOV-A on INSPIRE (16.5 pp). *Alt text: Two-panel figure visualizing the Simpson’s paradox gap across the eight external-validation runs. Top panel: paired bars per model, with each model’s overall AUC (low-saturation fill) shown beside the mean within-stratum AUC (hatched fill). For most preoperative models the within-stratum bar is markedly shorter than the overall bar, indicating loss of discrimination after stratifying by ASA. Asterisks flag the four INSPIRE-trained models on MOVER whose ASA 1–2 stratum has only nine events, so the within-stratum value falls back to the ASA* ≥ 3 *stratum alone. Bottom panel: paradox-gap (overall minus within-stratum, in percentage points) per model, with 95% bootstrap confidence interval whiskers; bar color encodes training cohort (blue = INSPIRE, brown = MOVER). The largest gap is XGB-MOV-A on INSPIRE at +16*.*5 pp*.

Across all eight external validations, paradox gaps ranged from 5.0 to 16.5 percentage points, with 95% bootstrap confidence intervals excluding zero in 8 of 8 models. Preoperative models showed systematically larger gaps than intraoperative models (mean 11.8 vs 6.8 pp; ratio 1.73). Bootstrap confidence intervals were tight (typical width 1.5–3.0 pp) and reproducible across resampling procedures.

Simpson’s paradox arises because preoperative models achieve overall discrimination primarily by separating patients into ASA risk strata, which have markedly different population-level mortality rates; when ASA’s contribution is removed through stratification, preoperative models provide minimal additional prognostic value within each stratum. Aggregate metrics therefore systematically overstate clinical utility when models separate strata without discriminating within them. We extend this analysis to cross-population transfer in §2.4.

### 2.4 Direction asymmetry: transferability depends on training population beyond measured case-mix

External validation revealed pronounced and statistically robust asymmetry in transfer direction (Table 3, Figure 3). INSPIRE-trained models showed mean external AUC degradation of 5.4 pp vs MOVER-trained models’ 13.9 pp — a 2.6-fold difference quantified by case-level paired bootstrap (2,000 iterations) at +8.53 pp (95% CI: +6.91–+10.24 pp; bootstrap p=0.001). All four MOVER-trained models showed greater external degradation than any of the four INSPIRE-trained models.

**Table 3:** Per-model direction asymmetry. Columns: training cohort, internal 10-fold-CV out-of-fold concatenated AUC, external AUC (after applying the trained model to the held-out alternate cohort), relative degradation (internal − external, pp; positive values indicate AUC loss on external transfer). The four MOVER-trained models all show greater external degradation than any of the four INSPIRE-trained models: a 2.6-fold mean difference.

| Model | Trained on | Internal AUC | External AUC | Degradation (pp) |
| --- | --- | --- | --- | --- |
| XGB-INS-A | INSPIRE | 0.807 | 0.785 | +2.7 |
| XGB-INS-B | INSPIRE | 0.860 | 0.895 | –4.1 |
| LR-INS-A | INSPIRE | 0.851 | 0.796 | +6.5 |
| LR-INS-B | INSPIRE | 0.887 | 0.741 | +16.4 |
| *INSPIRE-trained mean* |  | 0.851 | 0.804 | +5.4 |
| XGB-MOV-A | MOVER | 0.948 | 0.756 | +20.2 |
| XGB-MOV-B | MOVER | 0.942 | 0.812 | +13.7 |
| LR-MOV-A | MOVER | 0.914 | 0.806 | +11.8 |
| LR-MOV-B | MOVER | 0.931 | 0.839 | +9.9 |
| *MOVER-trained mean* |  | 0.934 | 0.804 | +13.9 |

**Figure 3:**
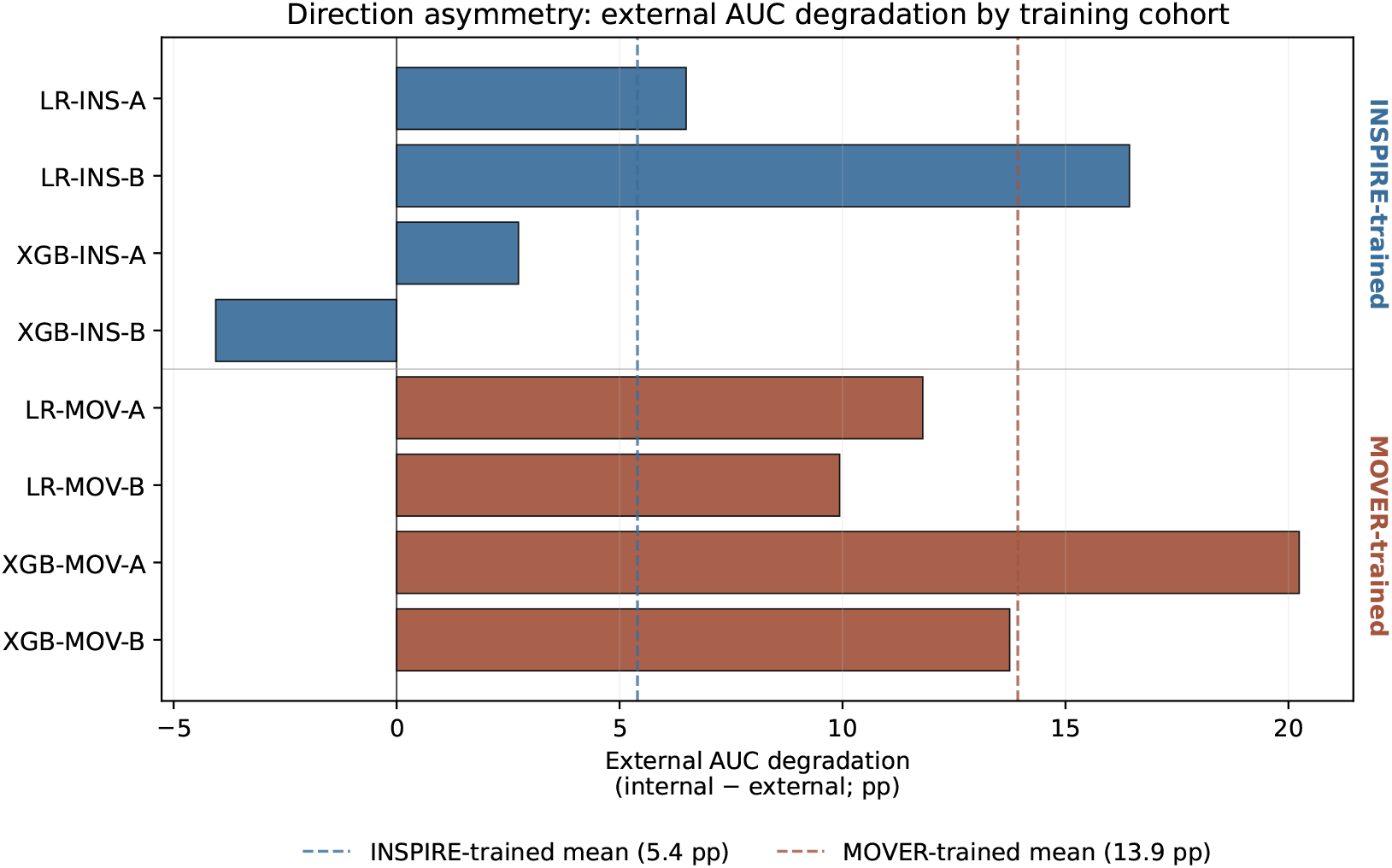
Direction asymmetry of external AUC degradation. Each bar is one model’s (internal AUC − external AUC); models grouped by training cohort. Dashed vertical lines mark within-group means. INSPIRE-trained models degrade by mean 5.4 pp; MOVER-trained models degrade by mean 13.9 pp, a 2.6-fold difference. All four MOVER-trained models exceed all four INSPIRE-trained models in degradation magnitude. *Alt text: Horizontal-bar chart of per-model external AUC degradation (internal AUC minus external AUC), grouped into two clusters by training cohort. The four INSPIRE-trained models are clustered at the top with shorter bars (smaller degradation, mean* ≈ *0*.*04 pp). The four MOVER-trained models are clustered at the bottom with longer bars (larger degradation, mean* ≈ *+8*.*6 pp), a roughly 2*.*6-fold difference. Dashed vertical lines mark each cluster’s mean. The visualization makes immediately visible that all four MOVER-trained models exceed all four INSPIRE-trained models in degradation magnitude — i*.*e*., *training direction matters beyond what within-cluster variation accounts for. Bar color matches Fig 2’s cohort encoding*.

#### Case-mix sensitivity across testable dimensions

To localize contributors, we conducted matched-subsampling across four case-mix dimensions (ASA stratum, Elixhauser comorbidity burden, emergency-case proportion, temporal period; Methods §3.4, full per-framing results in Supplementary Table S5). Temporal period matching was infeasible (INSPIRE timestamps are privacy-preserving relative-time offsets without a calendar-date mapping). For the three testable dimensions, three subsampling framings were applied (matching to each cohort’s distribution, A1 and A2; balanced target, B).

##### *ASA stratum matching* (not a primary driver)

Across three subsampling framings (matching to INSPIRE’s, MOVER’s, or a balanced distribution), matched asymmetries were 128–203% of the unmatched baseline (bootstrap *p* = 0.001 throughout). The pattern is mechanistically informative: INSPIRE-trained models matched to INSPIRE’s low-acuity distribution achieved external AUC exceeding internal AUC (degradation −3.4 pp; 95% CI: −7.4 to +1.3 pp), while MOVER-trained models matched to MOVER’s high-acuity distribution showed greater-than-baseline degradation (+17.0 pp; 95% CI: +15.5 to +18.4 pp).

##### *Elixhauser comorbidity matching* (not a primary driver)

Three framings produced asymmetries of 113–126% of baseline. MOVER’s higher comorbidity burden (mean van Walraven 5.94 vs INSPIRE’s 3.00) is consistent with concentration in ASA ≥ 3 patients but does not act as a hidden confounder.

##### *Emergency-case proportion matching* (partial contributor)

Matching INSPIRE up to MOVER’s 15.6% emergency rate produced +6.01 pp (95% CI: +4.25–+7.93 pp; bootstrap p=0.001). Across this and the reverse/balanced framings (MOVER to INSPIRE’s 7.9%; midpoint 11.75%), matched asymmetries fell to 70–86% of the unmatched baseline. Emergency-case burden contributes partially, with maximum ∼ 30% attenuation.

##### Consolidated interpretation (residual asymmetry)

Of the three testable dimensions, ASA stratum and comorbidity burden do not drive the asymmetry; emergency-case proportion is a partial contributor accounting for up to 30%. The residual asymmetry surviving matching on three dimensions — approximately 70% of the unmatched baseline, retaining bootstrap *p* = 0.001 — is attributable to training-population characteristics beyond measured case-mix, plausibly including healthcare-system-specific feature–outcome relationships, clinical-practice conventions, and unmeasured temporal-period effects (the fourth, untestable dimension; see Discussion §4.3).

#### Connection to Simpson’s paradox

Both findings share a common mechanism: preoperative models achieve apparent discrimination via stratum separation, easier on concentrated populations (MOVER’s 64% ASA ≥ 3) than spectrum-distributed populations (INSPIRE’s 90% ASA 1–2). The learned rule transfers poorly from concentrated to spectrum-distributed populations, contributing to MOVER-to-INSPIRE degradation; both phenomena require stratified, multi-dimensional analysis to detect.

### 2.5 Intraoperative features provide discrimination that transfers across populations, conditional on algorithmic capacity

A related question is whether specific feature classes — particularly intraoperative vital signs — transfer across populations. Mean intraoperative-feature AUC across the eight evaluations (0.822) exceeded mean preoperative-only AUC (0.786) by +3.60 pp (95% CI: +2.75 to +4.39 pp; bootstrap p=0.001). DeLong’s test with Benjamini–Hochberg correction confirmed significant intraoperative advantage in 3 of 4 within-direction comparisons: XGB-INS-B over XGB-INS-A (+11.0 pp, 95% CI +9.77 to +12.16; *p*_BH_ *<* 10^−16^), XGB-MOV-B over XGB-MOV-A (+5.6 pp, +4.51 to +6.85), and LR-MOV-B over LR-MOV-A (+3.3 pp, +2.63 to +3.93; both *p*_BH_ *<* 10^−16^).

The LR-INS-B exception describes a deployment failure mode that aggregate-statistic reporting masks. Logistic regression on the full 140-feature intraoperative set on the INSPIRE training direction (1,387 events; EPV = 9.9) underperformed its 8-feature preoperative counterpart on external validation by −5.5 pp (95% CI −7.84 to −3.17; *p*_BH_ = 4.6 × 10^−6^); XGBoost on the same direction showed no such degradation [21], its native L1/L2 regularization and early stopping protecting against overfitting under sparse-event conditions in a way standard logistic regression cannot. Where logistic regression is preferred for interpretability or in resource-constrained settings, high-dimensional features therefore require explicit regularization or pre-selection; this risk — algorithm choice interacting with feature-set size — likely generalizes beyond perioperative mortality, and is detectable only when per-model results are reported separately for each transfer direction. We retain LR-INS-B in all aggregate statistics; excluding it would raise the mean intraoperative advantage to approximately +5.8 pp.

### 2.6 Feature transfer: what transfers and what does not

A third question is *what* models are learning and whether learned feature relationships themselves transfer; SHAP feature-importance analysis and calibration evaluation together answer it.

#### Feature importance transfers near-perfectly across cohorts

Per-case SHAP values were computed using `shap.TreeExplainer` on the full external test set for the best model in each direction; rankings were compared between internal and external evaluations using Spearman rank correlation across all 140 shared features. Spearman correlations were *ρ* = 0.972 (XGB-INS-B, INSPIRE → MOVER) and *ρ* = 0.931 (XGB-MOV-B, MOVER → INSPIRE). The top feature was ASA in both directions, followed by BMI, age, and heart-rate-derived intraoperative features (Table S7; full SHAP summary plots in Supplementary Figures S4, S5), consistent with preoperative risk stratification via ASA and real-time physiological stability via HR-based intraoperative features as the primary discriminative axes in both cohorts.

#### Calibration does not transfer

Pre-recalibration calibration slopes ranged from 0.41 to 1.29 across the eight directions, and observed-to-expected (O:E) ratios were direction-asymmetric in a pattern that mirrors the §2.4 direction asymmetry. INSPIRE-trained models on MOVER systematically under-predicted mortality (O:E 0.02 to 0.10 across the four models), while MOVER-trained models on INSPIRE either modestly under-predicted (LR variants: O:E 0.10 to 0.19) or dramatically over-predicted (XGB variants: O:E up to 2.92); full per-model values are in Supplementary §S5. Platt scaling via 5-fold stratified cross-validation on the external dataset (each case held out of its own calibrator fit) restored calibration uniformly (post-recal slopes 0.95–1.02, O:E 0.99–1.01, mean Brier improvement ∼ 60%). AUC was rank-invariant under Platt scaling, so recalibration affected calibration and Brier but not discrimination. Table 4 reports the side-by-side preoperative-vs-intraoperative calibration values per model pair.

**Table 4:**
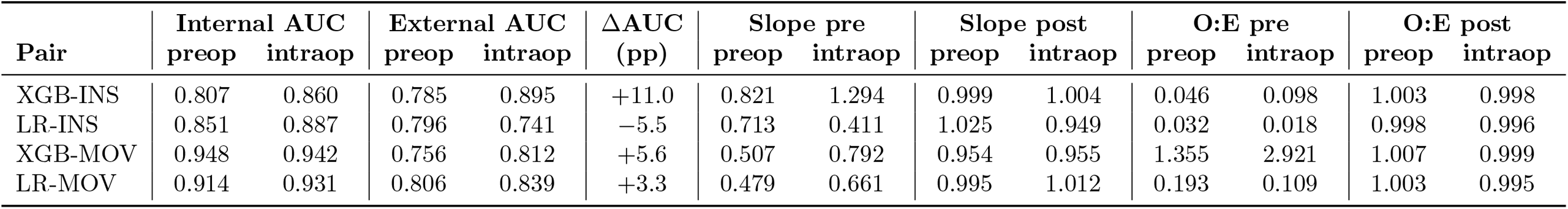
Side-by-side preoperative (A) vs intraoperative (B) discrimination and calibration per model pair. Each row reports an algorithm × training-cohort pair; columns alternate preop / intraop within each metric group. Internal AUC is the 10-fold OOF concatenated estimate; external AUC is the held-out alternate cohort. Calibration metrics are slope (slope = 1.0 ideal) and O:E (= 1.0 ideal), reported pre and post 5-fold cross-fit Platt scaling. The “ΔAUC intraop − preop” column quantifies the per-direction intraoperative-feature advantage; full per-model and per-stratum breakdowns are in Supplementary §S5 (Table S6).

### 2.7 Robustness

The four principal findings (§2.3–§2.6) describe how cross-population discrimination behaves; we next examine whether the findings themselves are robust to choices made during analysis. We tested robustness along five dimensions: demographic subgroup performance, model training configuration, alternative inferential frameworks, decision-theoretic clinical utility, and analyses constrained by data availability. The four-dimension case-mix sensitivity of the direction asymmetry finding (§2.4) is the most substantive robustness analysis in this paper. It is embedded in §2.4 rather than reported here because it localizes mechanism — identifying which case-mix dimensions contribute to the observed asymmetry — rather than stress-testing the headline claim. This section reports robustness checks applied uniformly across the principal findings. Full results appear in Supplementary Tables S10–S11.

#### Sex-stratified performance

Sex-stratified external AUC was computed for all eight models. Absolute female-versus-male AUC differentials ranged from 0.3 pp (XGB-INS-B on MOVER) to 3.4 pp (XGB-MOV-B on INSPIRE) across the eight models, with median |*F* − *M* | differential of 0.8 pp. We pre-specified a 5-percentage-point threshold for flagging fairness-relevant subgroup gaps, consistent with subgroup-audit practice in the clinical machine-learning fairness literature [18, 26]; no model exceeded this threshold in either direction. The largest observed differential (XGB-MOV-B on INSPIRE: female AUC 0.830 [95% CI 0.811–0.849] vs male AUC 0.796 [95% CI 0.780–0.811]; |Δ| = 3.4 pp) approaches but does not cross this threshold; models approaching this differential magnitude warrant sex-stratified AUC monitoring at deployment. The best-performing INSPIRE-trained model (XGB-INS-B) showed essentially no sex differential on MOVER (0.3 pp). The mild female advantage in MOVER-trained models applied to INSPIRE is consistent with the broader pattern that training-population diversity affects cross-population generalization.

#### Decision-curve clinical utility

Clinical utility was assessed using decision curve analysis across threshold probabilities of 0.5–29.5% (Figure 4). At clinically relevant thresholds (2–10% predicted mortality risk; chosen to span moderately above the 1.1–1.4% population base rate into clinically actionable decision ranges), recalibrated intraoperative models retained positive net benefit across the full 2–10% interval in three of four models (XGB-INS-B, XGB-MOV-B, LR-MOV-B); LR-INS-B lost utility above 2%, mirroring its within-direction degradation reported in §2.5. Among preoperative models, XGB-INS-A, XGB-MOV-A, and LR-MOV-A retained utility through the full 2–10% range, while LR-INS-A lost net-benefit advantage at thresholds above 7%. XGB-INS-B retained net-benefit advantage across the widest threshold range (0.5–29.5%). Intraoperative models showed a mean area under the precision-recall curve of 0.135 versus 0.091 for preoperative models (+49%), reflecting superior performance for rare-event prediction.

**Figure 4:**
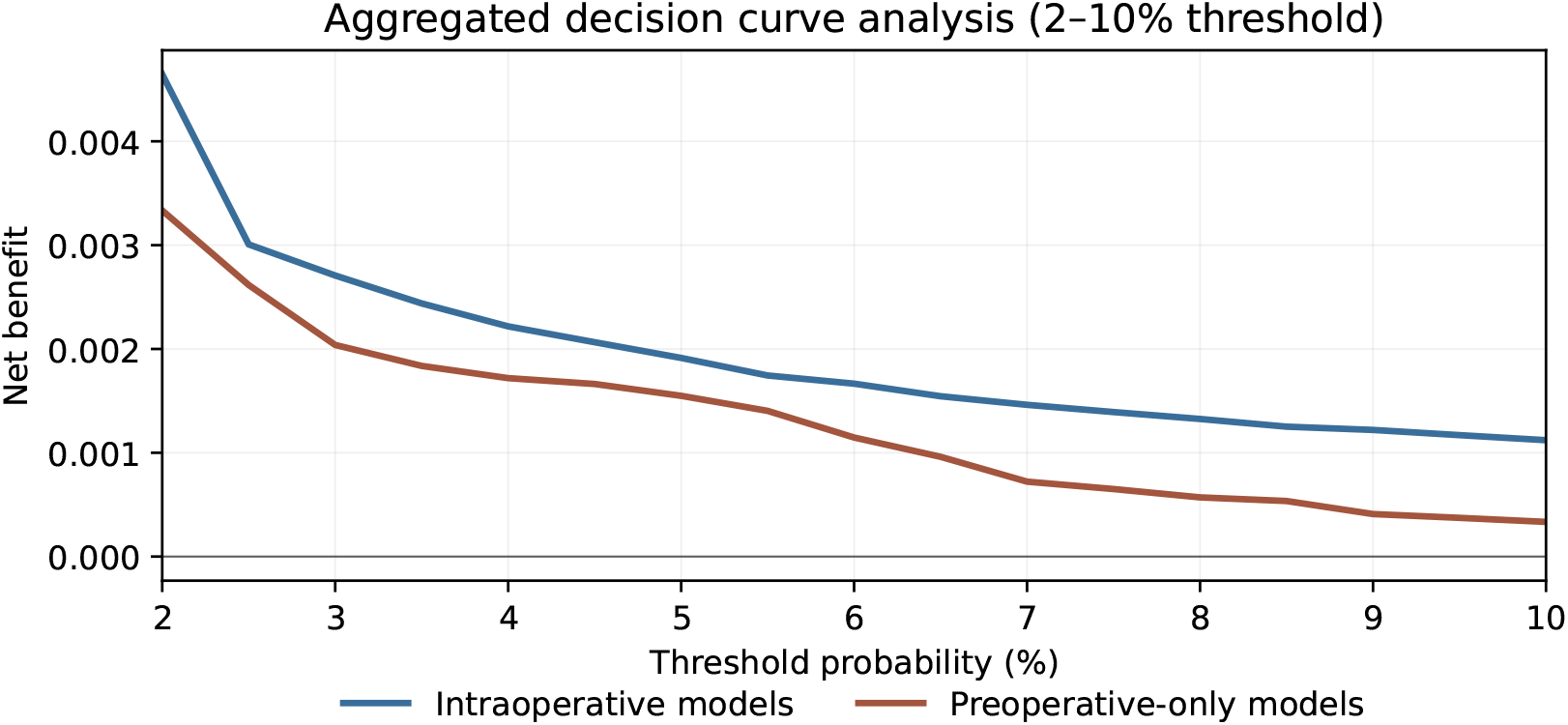
Decision curve analysis at clinically relevant 2–10% threshold range. Mean net benefit is shown for the four intraoperative-feature (B) models (blue) and the four preoperative-only (A) models (brown), each averaged across the two transfer directions. Treat-all reference is in Supplementary Figure S6. Recalibrated intraoperative models retain positive net benefit across the 2–10% interval; per-direction breakdown showing the LR-INS-B utility-loss above 2% is in Supplementary Figure S6. *Alt text: Decision-curve analysis plot showing mean net benefit (vertical axis) as a function of decision threshold probability (horizontal axis, 2% to 10%). Two curves are shown: (i) mean of the four intraoperative-feature models in blue, and (ii) mean of the four preoperative-only models in brown, each averaged across the two transfer directions (INSPIRE*→*MOVER and MOVER*→*INSPIRE). The intraoperative-feature curve sits above the preoperative-only curve across the entire 2–10% threshold range, indicating that recalibrated intraoperative models retain positive net benefit for clinically realistic mortality decision thresholds. The plot’s “treat-all” reference line (constant net benefit assuming all patients are treated) is shown in the per-direction supplementary breakdown rather than this main aggregate panel*.

#### Class imbalance

Three configurations of XGB-INS-B were compared: no class weighting, the representative class weighting (*s*_*pos*_ = 9.73, the median per-fold tuned value from the main analysis described in §3), and SMOTE oversampling. Discrimination was comparable between no weighting (AUC 0.898) and the representative class weighting (AUC 0.895); SMOTE reduced external AUC by 7.7 pp (AUC 0.822), consistent with recent evidence that synthetic oversampling harms calibration and discrimination in rare-event clinical prediction [3].

Inferential robustness across alternative resampling configurations and three additional analyses constrained by data limitations are reported in Supplementary §S5.

## 3 Materials and Methods

### 3.1 Datasets and cohort

Two publicly available perioperative datasets were used: INSPIRE [15] from Seoul National University Hospital (Korea; 2011–2020) and MOVER [23] from UC Irvine Medical Center (USA; 2015–2022). The primary outcome was in-hospital mortality, defined for INSPIRE as a non-null `inhosp_death_time` timestamp and for MOVER as a discharge disposition of `Expired` or `Coroner`. Final cohort sizes after inclusion and feature-completeness filtering: 127,413 (INSPIRE) and 57,545 (MOVER).

Inclusion required a recorded preoperative ASA physical status (1–6), age, and sex. MOVER’s source release implicitly excluded cardiac-catheterization-laboratory and interventional-radiology procedures (no ASA assignment); INSPIRE had no analogous exclusion. Records without a recorded mortality outcome at hospital discharge were excluded from MOVER (*n* = 3 of 58,758). Per-cohort criteria are detailed in Supplementary §S1; the CONSORT flow is shown in Figure 1.

Both datasets were accessed under their respective data-use agreements; no patient-level data are redistributed in this paper.

### 3.2 Models

Eight models were trained in a 2× 2× 2 factorial design: algorithm (XGBoost, logistic regression) × feature set (8-variable preoperative; 140-variable preoperative+intraoperative) × training dataset (INSPIRE, MOVER). Intraoperative features were summary statistics of vital signs (heart rate, blood pressure, oxygen saturation, end-tidal CO_2_, temperature) over the 60-minute post-induction window [11]. Hyperparameters were tuned by Bayesian optimization with 10-fold stratified cross-validation; class imbalance was addressed via XGBoost’s per-fold-tuned scale-positive weight, with SMOTE evaluated as a sensitivity analysis (§2.7) but not used in the main analysis [3]. Events-per-variable (EPV) ratios were 173 and 9.9 for INSPIRE preoperative and intraoperative models, and 103 and 5.9 for MOVER; the intraoperative EPV ratios approach the recommended lower threshold for stable logistic regression in rare-event settings [21], examined in §2.5. Full training procedures, hyperparameter search spaces, and feature definitions are in Supplementary §S2.

### 3.3 Statistical framework

Case-level paired bootstrap (2,000 iterations, `seed = 42`) served as the primary inferential framework for all quantitative comparisons reported in §2. Bootstrap samples were drawn with replacement from cases within each external test set; paired-bootstrap replicates were used for AUC differences and paradox gaps. Bootstrap p-values reported as *p* = 0.001 correspond to the Monte Carlo floor of 1*/B* for *B* = 2,000 resamples — zero of 2,000 replicates crossed the null. These values are upper bounds on the true tail probability rather than point estimates. The 4-vs-4 model-level permutation test has minimum achievable 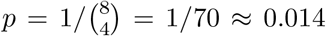 regardless of effect magnitude, motivating case-level (*n* = 127,413 and 57,545) rather than model-level (*n* = 8) inference for between-group comparisons. DeLong’s test [7] with Benjamini–Hochberg correction was applied to pairwise AUC comparisons as a robustness check. Post-hoc Platt scaling [20] was applied for recalibration on external datasets; AUC values are reported separately from calibration statistics because AUC is rank-invariant under monotonic recalibration. Full bootstrap procedure, AUC variance estimation, and correction procedures are in Supplementary §S3.

### 3.4 Matched-subsampling sensitivity for direction asymmetry

Cross-continental degradation asymmetry was stress-tested across four case-mix dimensions: ASA stratum, Elixhauser comorbidity (Van Walraven-weighted scores derived via the Quan ICD-10 mapping), emergency-case proportion, and temporal period. For each non-temporal dimension, three subsampling framings (matching each cohort’s distribution and a balanced midpoint) were applied; bootstrap CIs were computed using the same case-level paired bootstrap (2,000 iterations, seed 42) as the unmatched primary analysis. Temporal matching was not feasible because INSPIRE distributes timestamps as privacy-preserving relative-time offsets without published calendar-date mappings. Full pseudocode for the two-stratum maximum-*n* subsampling algorithm and worked examples are in Supplementary §S4.

### 3.5 Feature importance and calibration

SHAP values (`shap.TreeExplainer`) were computed on external test sets for the top XGBoost model in each direction (the B-model, using all 140 features); internal values were aggregated from per-fold held-out predictions, and rankings compared via Spearman rank correlation across all 140 shared features (features below the 25th percentile of mean absolute SHAP value were filtered as a noise floor for the transferable-features sub-analysis). Calibration was assessed via slope, intercept, observed-to-expected ratios, and Brier score before and after recalibration. Platt scaling was fit on the external dataset via 5-fold stratified cross-validation, each case held out of its own calibrator fit. Full procedures: Supplementary §S5.

### 3.6 Reporting and reproducibility

This study follows TRIPOD+AI reporting guidelines [6] (checklist: Supplementary Table S1). Code is archived at Zenodo (DOI: 10.5281/zenodo.19941764, concept DOI resolving to the latest version) under an MIT license; trained XGBoost and logistic-regression artifacts are deposited alongside, pending steward confirmation that the INSPIRE (PhysioNet) and MOVER (UC Irvine) data-use agreements permit open-repository model-weight release. We interpret both DUAs conservatively to prohibit redistribution of patient-level derivatives, including per-case prediction outputs; credentialed researchers can reproduce all analyses by applying the released models to the source data. Aggregated statistics covering every main-text claim are provided in Supplementary §S6, subject to each DUA’s terms and any minimum-cell-size requirements. Software dependencies are pinned and random seeds fixed.

## 4 Discussion

### 4.1 Principal findings

Aggregate AUC reporting concealed within-stratum AUCs near the no-discrimination floor, with paradox gaps to 16.5 pp in the worst-affected preoperative model. Cross-population transferability ran 8.5 pp better in one direction than the other, persisted at roughly 70% of baseline after multi-dimensional case-mix matching, and was invisible to any single-direction validation. Pre-Platt calibration slopes spanned 0.41–1.29 across all eight cross-population runs and recovered to 0.95–1.02 only after 5-fold cross-validated Platt scaling. Each of these magnitudes is clinically material at the patient-decision threshold and would not have surfaced under conventional aggregate evaluation. The methodological commitments that surface them are individually well-established TRIPOD+AI-aligned practices: stratified within-cohort analysis across clinically meaningful risk strata; bidirectional testing with multi-dimensional matched case-mix sensitivity; and case-level paired bootstrap inference with rank-invariant metrics reported separately from probability-scale metrics. The sections that follow position each finding against the literature (§4.2), examine the underlying mechanisms (§4.3), derive methodological implications (§4.4), and consider the constraints that limit our conclusions (§4.5).

### 4.2 Comparison with existing literature

Recent perioperative-medicine systematic review [2] reports that only 13% of published machine-learning prediction models undergo external validation, and that single-center development dominates the literature; PROBAST assessments identify high risk of bias in most models. Within this landscape, the magnitudes we report (paradox gaps to 16.5 pp, direction asymmetry of 8.5 pp persisting at ∼70% of baseline after matched-case-mix sensitivity, calibration slope ranges from 0.41 to 1.29 across cross-population runs) characterize what survives bidirectional testing and stratified evaluation in the rare case that both can be applied. To our knowledge, paradox-gap magnitudes of this scale have not previously been reported in the deployed clinical prediction model literature; Chipman and Braun [5] established the analytic basis for stratum-level miscalibration producing paradox in performance metrics, and our findings extend that pattern to AUC in cross-continental perioperative deployment.

The direction-asymmetry finding extends a smaller cross-population literature: Futoma et al. [12] and Finlayson et al. [10] characterized the “myth of generalisability” and distributional shift as core deployment risks but did not quantify direction-of-transfer asymmetry or provide a case-level paired bootstrap framework. The SHAP transferability finding (Spearman *ρ* = 0.93–0.97) characterizes feature-importance stability between geographically separated academic medical centers; prior SHAP-robustness work focused on within-dataset stability [16], and the cross-population result suggests that the relational structure of what models learn is more robust than the absolute calibration on probability scale.

### 4.3 Mechanisms

The four-dimension matched-sensitivity analysis clarifies the mechanism underlying the observed direction asymmetry: ASA stratum matching amplifies the asymmetry to 128–203% of baseline; Elixhauser comorbidity matching preserves or amplifies it (113–126%); emergency-case proportion attenuates it to 70–86% but does not eliminate it; and temporal period was untestable due to INSPIRE timestamp anonymization. The residual asymmetry surviving the three testable matches — approximately 70% of baseline — is attributable to training-population characteristics beyond measured case-mix: healthcare-system-specific feature–outcome relationships, within-stratum severity coding that ASA and Elixhauser do not capture, and unmeasured temporal effects. At matched ASA stratum, INSPIRE deaths spread across the acuity spectrum (47.2% / 52.8% in ASA 1–2 / ≥ 3) while MOVER deaths concentrate in high-acuity patients (1.1% / 98.9%) — a 50.6-percentage-point split-difference consistent with cross-institutional feature–outcome relationships differing beyond what stratum matching equalizes. ASA physical status itself shows substantial inter-rater variability across institutions and clinicians [25], so a “severity-3” patient under INSPIRE coding may not be clinically equivalent to a “severity-3” patient under MOVER coding. Cross-institutional surveys of Asian-ICU end-of-life decision-making have documented systematic differences between Korean and Western practice patterns [17, 19], consistent with the healthcare-system-specific mechanism interpretation above. Supplementary §S7 reports stratified mortality distributions and the supporting cross-institutional literature.

That ASA stratum, comorbidity burden, and emergency-case proportion all leave most of the asymmetry intact, and that ASA matching in fact amplifies it, is the strongest evidence we can offer that the finding is not an artifact of case-mix mismatch. The temporal dimension we could not test directly; INSPIRE’s privacy-preserving relative timestamps foreclose calendar-aligned matching. We name this gap because it sets a real boundary on confidence: the residual asymmetry could in principle reflect temporal-period effects we have no way to estimate. The finding stands; future work benefits from cohorts where calendar-date alignment is available.

The SHAP-plus-calibration finding provides a mechanistic lens on what transfers. Feature importance rankings are preserved almost perfectly (*ρ* ≥ 0.93): a Seoul-trained model evaluated on Irvine data ranks feature importance (ASA, BMI, HR dynamics) in the same order as on its training cohort, yet absolute risk probabilities are systematically biased and require recalibration. Calibration fails uniformly before Platt scaling (O:E 0.02 to 2.92; slopes 0.41 to 1.29). This dissociation — discrimination is preserved as a ranking, but calibration is distorted on the probability scale — aligns with the well-documented pattern that discrimination transfers more readily than calibration [28], supporting the interpretation that cross-population models preserve relational structure (which features matter, in what order) but not absolute mappings (what probability a given feature vector implies).

Returning to the asymmetry finding, this case study cannot directly answer how the pattern might run with training drawn from an LMIC tertiary perioperative cohort. Healthcare-system-specific feature–outcome relationships, named in the residual-mechanism analysis above as one component of the asymmetry that case-mix matching cannot equalize, are especially relevant in this context. LMIC tertiary perioperative care chains differ from HIC counterparts in ways case-mix variables do not capture: referral patterns, selection-into-surgery under limited non-operative alternatives, and post-operative monitoring infrastructure all shape feature–outcome relationships in systematically different ways. Whether models trained on LMIC perioperative cohorts would transfer to HIC deployment more or less successfully than the conventional HIC→LMIC direction is not something present data can settle; paired LMIC–HIC perioperative datasets at INSPIRE or MOVER scale do not yet exist. The asymmetry mechanism makes the question relevant for deployment contexts where consequences are largest.

### 4.4 Clinical implications

These findings motivate three concrete changes to current practice. First, stratified external validation within clinically meaningful risk groups (e.g., ASA 1–2 vs ≥ 3; elective vs emergency) should become standard for TRIPOD+AI-compliant reporting; aggregate AUC should accompany, not replace, within-stratum AUCs and paradox gaps (Supplementary Table S9). Second, cross-population claims should include bidirectional transfer testing with multi-dimensional matched-case-mix sensitivity; *A* → *B* without reciprocal *B* → *A* cannot detect direction asymmetry, and matching is required to distinguish genuine failure from case-mix mismatch. Third, cross-population deployment requires mandatory probability recalibration (Platt or isotonic) on a representative sample of the deployment population [9]. Recent external-validation sample-size guidance suggests that several thousand cases are typically needed to estimate calibration slope and O:E precisely, depending on the anticipated event proportion and degree of miscalibration [22]. Discrimination-focused reporting alone is insufficient; published AUCs should accompany calibration assessments and re-calibration protocols.

These methodological commitments are particularly consequential for HIC → LMIC deployment of clinical prediction models, where healthcare-system-specific feature–outcome relationships (§4.3) and the absence of paired LMIC–HIC perioperative datasets at scale together constrain the evidence base reviewers and deployers can draw on; the cautionary patterns we report (direction asymmetry, calibration collapse, paradox gaps) are precisely the failure modes such deployments face.

### 4.5 Limitations

Five principal limitations apply. First, only two institutions (Seoul National and UC Irvine) were included, characterizing cross-institutional rather than cross-national transfer. Second, temporal-period matching was infeasible due to INSPIRE’s privacy-preserving relative-time offsets; this constraint and its implications for the residual asymmetry are addressed in §4.3. Third, neither race/ethnicity stratification (omitted from the public MOVER release; no Korean comparator) nor within-stratum severity matching beyond Elixhauser comorbidity could be uniformly operationalized; race/ethnicity is therefore not analyzed. This prevents detection of subgroup discrimination failures along race or ethnicity — a well-documented fairness failure mode in cross-population deployment [4, 18]. Fourth, procedure-type matching was not attempted because INSPIRE’s KCD-10 and MOVER’s CPT/EPIC code taxonomies do not align without a purpose-built crosswalk. Fifth, the 60-minute post-induction window was chosen by clinical convention; alternative lengths were not systematically examined. These constraints bound the empirical reach of the present case study but do not affect its standing as a cautionary reference for cross-population evaluation practice.

## 5 Conclusion

This case study reports cross-continental perioperative mortality prediction failure modes at clinically material magnitudes. We position these findings as a cautionary reference for cross-population deployment of clinical prediction models, in the lineage of post-deployment evaluation work in clinical machine learning [29]. Trained model objects, analysis code, and aggregate verification outputs are archived for reproducibility (§3.6). Paired LMIC–HIC perioperative cohorts with calendar-aligned timestamps are a natural next step.

## Data Availability

INSPIRE is publicly available via PhysioNet under credentialed access (DOI 10.13026/46m4-f655); MOVER is publicly available via the UC Irvine Machine Learning Repository under credentialed access (DOI 10.24432/C5VS5G). Both datasets were accessed under their respective data-use agreements; no patient-level data are redistributed in this paper.
Analysis code, figure-generation scripts, trained model objects, and aggregate analysis outputs are archived at Zenodo (DOI 10.5281/zenodo.19941764; concept DOI resolving to the latest version; see Methods 3.6).
Per-case prediction outputs are not redistributed (data-use agreement constraint); credentialed researchers can reproduce all analyses by applying the released models to source data after re-credentialing on PhysioNet and the UCI Machine Learning Repository.

https://physionet.org/content/inspire/1.3/

https://archive.ics.uci.edu/dataset/877/mover-medical-informatics-operating-room-vitals-and-events-repository

https://doi.org/10.5281/zenodo.19941764

## Acknowledgments

The author acknowledges the Seoul National University Hospital INSPIRE team for stewardship of the INSPIRE dataset [15] and the UC Irvine perioperative team for stewardship of the MOVER dataset [23].

## Author Contributions

Sole author. The author was responsible for all aspects of the work, including study conception and design, data acquisition under credentialed-access agreements, software implementation, formal analysis, interpretation, manuscript drafting, and final approval. The CRediT taxonomy roles — conceptualization, methodology, software, validation, formal analysis, investigation, resources, data curation, writing (original draft), writing (review and editing), visualization, supervision, project administration, and funding acquisition — all apply to the sole author.

## AI Use Disclosure

Anthropic’s Claude (Claude Sonnet 4 and Claude Opus 4 series, accessed via Claude Code and the Claude.ai web interface, 2025–2026) was used during preparation of this manuscript for two purposes: (i) generation and refinement of analysis and figure-generation code in Python, which was reviewed and validated by the author before use; and (ii) editorial assistance with prose revision, copyediting, and structural organization of author-written drafts. All scientific claims, statistical analyses, study design choices, and interpretations are the author’s; every quantitative result reported was independently validated against source data and primary literature by the author. No AI system is listed as an author, consistent with COPE and ICMJE guidance that AI tools cannot meet authorship criteria.

## Data Availability Statement

INSPIRE is publicly available via PhysioNet under credentialed access [14]; MOVER is publicly available via the UC Irvine Machine Learning Repository under credentialed access [24]. Both datasets were accessed under their respective data-use agreements; no patient-level data are re-distributed in this paper. Analysis code, figure-generation scripts, trained model objects, and aggregate analysis outputs are archived at Zenodo (DOI: 10.5281/zenodo.19941764; see Methods §3.6). Per-case prediction outputs are not redistributed (data-use agreement constraint); credentialed researchers can reproduce all analyses by applying the released models to source data after re-credentialing on PhysioNet and the UCI Machine Learning Repository.

## Ethics Statement

This study used de-identified data released for research use under the data-use agreements of INSPIRE (PhysioNet) and MOVER (UC Irvine Machine Learning Repository). No additional Institutional Review Board review was sought, as the work involves only secondary analysis of publicly released, de-identified datasets accessed under credentialed-use agreements. The author followed the data-use terms of both repositories.

