## Supplementary Material for "When external validation isn’t enough: Simpson’s paradox, direction asymmetry, and calibration collapse in cross-continental perioperative mortality prediction"

**S8 TRIPOD+AI reporting checklist** **21**

### S1 Cohort definition and inclusion/exclusion

This section documents per-cohort inclusion and exclusion criteria with the case counts at each filter stage. The primary outcome and EPV reasoning are defined in main Methods §3.1; mortality counts, rates and per-stratum breakdowns are reported in main Results §2.1 and verified in §S6. Both cohorts were accessed under credentialed agreements (PhysioNet for INSPIRE; UC Irvine for MOVER); no patient-level data were redistributed.

#### S1.1 INSPIRE cohort

The INSPIRE source release is the v1.3 PhysioNet deposit [7] (Seoul National University Hospital, 2011–2020). Of the 130,960 operations in the source release, all records carried valid subject and operation identifiers; 3,547 were excluded for missing or out-of-range ASA classification (the only inclusion-related attrition step), and no further records were excluded for missing age or sex (both fields are complete in the records that pass ASA filtering). The final analysis cohort is 127,413 surgical encounters. The primary outcome (in-hospital mortality from a non-null `inhosp_death_time` timestamp; main Methods §3.1) was ascertainable for all 127,413 cases. The feature-completeness step introduces no further attrition from the cohort-definition step described above, and intraoperative-feature coverage in the 60-minute post-induction window is 99.9% (the 0.1% missing cases retain preoperative-feature predictions and are imputed downstream).

#### S1.2 MOVER cohort

The MOVER source release is the UC Irvine Medical Center EPIC subset [12] (2015–2022). Of 65,728 surgeries in the source release, all records had valid `MRN` and `LOG_ID`; 6,970 were excluded for missing ASA classification (these are predominantly cardiac-catheterization-laboratory and interventional-radiology procedures, which receive no anesthesiologist ASA assignment), leaving 58,758. A further 3 records were excluded for missing discharge disposition (the outcome-ascertainment field), leaving 58,755. At a downstream deduplication step, an additional 1,210 records were dropped as duplicate (MRN, LOG_ID) pairs introduced by an upstream emergency-flag left-merge step (deduplicated by `keep="first"` on first occurrence). The final analysis cohort is 57,545 surgical encounters. The primary outcome (`DISCH_DISP` $\in$ {Expired, Coroner}; main Methods §3.1) was ascertainable for all 57,545 cases; intraoperative-feature coverage in the 60-minute post-induction window is 92.1%, with the gap attributable to records lacking `AN_START_DATETIME`.

Table S1: Per-cohort inclusion/exclusion attrition counts. "—" marks a filter stage that is not applicable to the cohort (INSPIRE has no discharge-disposition or post-merge-dedup attrition because no equivalent upstream merge step was used).

| **Step** | **INSPIRE** $n$ | **MOVER** $n$ |
|---|---|---|
| Source data | 130,960 | 65,728 |
| After: ASA 1–6 recorded | 127,413 | 58,758 |
| After: discharge disposition non-null | — | 58,755 |
| After: age and sex non-null | 127,413 | 58,755 |
| After: (MRN, LOG_ID) deduplication | — | 57,545 |
| **Final analysis cohort** | **127,413** | **57,545** |

### S2 Model training

This section documents the feature schema, hyperparameter search spaces, median tuned values, and class-imbalance protocol used to train all eight models. The high-level architecture and EPV reasoning live in main Methods §3.2; §S2 is the implementation appendix.

#### S2.1 Feature definitions

The four **preoperative-only (A) models** use 8 features extracted from the harmonized cohort tables: `age`, `sex`, `height_cm`, `weight_kg`, `bmi`, `asa`, `emergency`, and the derived stratification indicator `high_asa` (8 features in total). The four **preoperative+intraoperative (B) models** use these 8 plus 132 additional features for a total of 140: 3 categorical/derived contextual features (`anesthesia_type`, `department`, `high_asa_emergency`) and 129 intraoperative vital-sign summary statistics extracted from the 60-minute post-induction window [6]. Vital signs included heart rate (HR), pulse oximetry ($SpO_2$), respiratory rate (RR), temperature, non-invasive systolic/diastolic blood pressure (SBP, DBP), mean arterial pressure (MAP, with arterial-line variants where available), end-tidal $CO_2$ ($EtCO_2$), and inspired $O_2$ fraction ($FiO_2$); each yielded mean/min/max/SD/first/last summaries plus threshold-time features (e.g., `hr_below_50`, `temp_below_35`, `temp_above_38`). $EtCO_2$ coverage in INSPIRE was verified at 93.93%. Note: "140 features" in main Methods §3.2 and "132 intraoperative features" here refer to the same intraoperative-feature set with different counting conventions (140 includes the 8 preoperative features; 132 excludes them).

Table S2: Feature inventory for the 8-feature preoperative-only (A) and 140-feature preoperative+intraoperative (B) model families.

| **Group** | **Count** | **In A?** | **Examples** |
|---|---|---|---|
| Preoperative | 8 | ✓ | age, sex, BMI, ASA, emergency, high_asa |
| Contextual (B-only) | 3 | | anesthesia_type, department |
| Vital-sign summaries (B-only) | 129 | | hr_mean, map_min, sbp_std, etco2_first |
| of which threshold features | (subset) | | hr_below_50, temp_above_38 |
| **B-model total** | **140** | | |

#### S2.2 Hyperparameter search and tuning

XGBoost hyperparameters were tuned via Bayesian optimization (Tree-structured Parzen estimator via Optuna [1]; 50 trials per outer fold) within a 10×5 nested cross-validation framework: 10 stratified outer folds for unbiased estimation and 5 inner folds for HPO. The search space (Table S3) was code-verified against the training infrastructure source. `n_estimators` was excluded from the search (capped at 2,000 with early stopping at 50 rounds without improvement on validation AUC). The HPO eval metric was AUC; Platt scaling (§S3.5) was used for calibration. The `scale_pos_weight` range was set per-fold to $[1.0, n_{\text{neg}}/n_{\text{pos}}]$, giving an upper bound of $\approx 91$ for INSPIRE folds and $\approx 69$ for MOVER folds. Logistic regression used a coarse grid over the inverse regularization strength $C \in \{10^{-3}, 10^{-2}, 10^{-1}, 1, 10, 10^2\}$ with L2 penalty, `class_weight=balanced`, `lbfgs` solver, and `max_iter=1,000`; missing values were imputed via `SimpleImputer` (mean) before standardization. Median tuned hyperparameters across the 10 outer folds are reported per model in Table S4. The XGB-INS-B median `scale_pos_weight` of 9.73 is the value used as the representative single-value setting in the class-imbalance ablation (§S2.3 and main §2.7).

Table S3: XGBoost hyperparameter search space. Distribution column gives the Optuna TPE sampler distribution used for each parameter.

| Parameter | Range | Distribution |
|---|---|---|
| max_depth | $[2, 10]$ | uniform integer |
| learning_rate | $[10^{-3}, 0.3]$ | log-uniform |
| min_child_weight | $[1, 20]$ | uniform integer |
| gamma | $[0.0, 5.0]$ | uniform |
| reg_alpha | $[10^{-8}, 1.0]$ | log-uniform |
| reg_lambda | $[10^{-8}, 10.0]$ | log-uniform |
| subsample | $[0.5, 1.0]$ | uniform |
| colsample_bytree | $[0.5, 1.0]$ | uniform |
| scale_pos_weight | $[1.0, n_{\text{neg}}/n_{\text{pos}}]$ per fold | uniform (per-fold-dynamic) |
| n_estimators | not searched (max 2,000; early stop at 50) | — |

Table S4: Median tuned XGBoost hyperparameters across the 10 outer folds, per model. `spw` = `scale_pos_weight`; min/max columns show the per-fold range. `T` = `n_estimators` chosen by early stopping (median across folds; capped at 2,000, early-stop patience 50).

| Model | depth | lr | mcw | sub | cs | $\gamma$ | $\alpha$ | $\lambda$ | spw med | spw range | T |
|---|---|---|---|---|---|---|---|---|---|---|---|
| XGB-INS-A | 4 | 0.062 | 8 | 0.87 | 0.73 | 2.64 | 5.6e-6 | 7.7e-5 | 21.79 | 3.25–65.22 | 99 |
| XGB-INS-B | 4 | 0.014 | 9 | 0.69 | 0.81 | 2.92 | 3.4e-4 | 4.7e-5 | 9.73 | 1.84–67.39 | 464 |
| XGB-MOV-A | 10 | 0.073 | 5 | 0.64 | 0.62 | 2.75 | 4.2e-5 | 2.1e-3 | 41.23 | 5.51–67.44 | 164 |
| XGB-MOV-B | 10 | 0.039 | 7 | 0.76 | 0.79 | 2.69 | 8.2e-4 | 7.2e-4 | 15.58 | 4.66–50.03 | 355 |

##### S2.3 Class imbalance handling

Three distinct class-imbalance protocols appear in this study and are documented here to avoid conflation. **(i) Main analysis (XGBoost):** `scale_pos_weight` was tuned independently per outer fold over $[1.0, n_{\text{neg}}/n_{\text{pos}}]$ by the TPE sampler; the resulting per-fold values span the ranges shown in Table S4 (e.g., XGB-INS-B folds ranged 1.84 to 67.39). **(ii) Main analysis (LR):** `class_weight="balanced"` was used (sklearn's inverse-frequency weighting), held fixed across folds. **(iii) Class-imbalance ablation (XGB-INS-B only; main §2.7):** to isolate the imbalance-treatment effect from hyperparameter variation, three configurations of XGB-INS-B were retrained with all hyperparameters fixed at the median values from Table S4: no weighting (`scale_pos_weight = 1`); class weighting (`scale_pos_weight = 9.734`, the median of the 10 per-fold tuned values); and SMOTE oversampling (minority class oversampled to parity with `k_neighbors=5`, applied to the training fold only, `scale_pos_weight = 1`). The SMOTE result (7.7-pp external AUC reduction vs. no weighting) is reported in main §2.7 and is consistent with prior evidence [2].

#### S3 Statistical framework

This section documents the unified inferential machinery used for every quantitative claim in main Results: case-level paired bootstrap (the primary framework), the Monte Carlo floor reporting convention, the model-level permutation test (sensitivity), DeLong's test with Benjamini–Hochberg correction (per-pair sensitivity), and the Platt 5-fold cross-validation procedure (recalibration). Pseudocode is code-verified against the analytical pipeline.

#### S3.1 Case-level paired bootstrap

The primitive shared across every inferential claim in this study is a case-level paired bootstrap with $B = 2{,}000$ iterations and `seed = 42`. For each iteration, an index vector of length $n_{\text{group}}$ is drawn with replacement *independently per cohort group* (separately for the INSPIRE test set and the MOVER test set; never across cohorts, both because the two cohorts are independent draws and because the credentialed-data DUAs prohibit case-level mixing). Within an iteration, every statistic computed on a given test set re-uses the same index vector, which is what makes the bootstrap *paired*: differences between models on the same external set have correlated bootstrap error, so differences-of-differences are well-estimated. Percentile 95% CIs are reported. Two-sided bootstrap p-values are computed as $p = 2 \times \min(\Pr[\hat{\theta}^* \leq 0], \Pr[\hat{\theta}^* \geq 0])$ with $+1/+1$ continuity correction.

**Algorithm 1** Case-level paired bootstrap (per cohort).

**Require:** Per-group case counts $\{n_g\}$; statistic function $T(\cdot)$; $B = 2{,}000$; $\texttt{seed} = 42$
**Ensure:** 95% percentile CI and two-sided $p$ for each statistic

```
Initialize RNG with seed; allocate empty arrays T_b
for b = 1, ..., B do
    for each group g do
        I_g ← RNG.INTEGERS(0, n_g, n_g)                    ▷ draw with replacement
    end for
    T_b ← T({I_g})                                         ▷ statistic on resampled cases
end for
Sort T_b; report T̂ = median(T_b), CI_95% = [P_2.5, P_97.5]
p ← 2 · min(#{T_b ≤ 0} + 1, #{T_b ≥ 0} + 1) / (B + 1)
return T̂, CI_95%, p
```

#### S3.2 Monte Carlo floor and p-value reporting convention

With $B = 2{,}000$ replicates the smallest non-zero bootstrap p-value estimable is $1/B = 5 \times 10^{-4}$. A bootstrap with zero of 2,000 replicates crossing the null is reported throughout main text, abstract, and this supplementary as $p = 0.001$ (with the $+1/+1$ continuity correction this reads $2/(B+1) \approx 1.0 \times 10^{-3}$). This convention is consistent with the convention used in the canonical Methods §3.3: $p = 0.001$ is an *upper bound* on the true tail probability, not a point estimate.

#### S3.3 Model-level permutation test

The 4-vs-4 model-level permutation test (a sensitivity check for the direction-asymmetry claim) has minimum achievable $p = 1/\binom{8}{4} = 1/70 \approx 0.014$ regardless of effect magnitude, because the maximum number of distinct re-labelings of 8 model labels into 2 groups of 4 is 70. This bound is independent of how separated the two groups are and is the reason main Methods §3.3 motivates case-level (n = 127,413, 57,545) rather than model-level (n = 8) inference for between- group comparisons. The case-level paired bootstrap (§S3.1) on the same data yields $p = 0.001$ for the same direction-asymmetry claim.

#### S3.4 DeLong's test with Benjamini–Hochberg correction

DeLong et al. [5] provide a nonparametric significance test for differences between paired AUCs. The test was applied here as a robustness check on the four within-direction preop-vs-intraop pairs reported in main §2.5: XGB-INS-B vs XGB-INS-A, LR-INS-B vs LR-INS-A, XGB-MOV-B vs XGB-MOV-A, and LR-MOV-B vs LR-MOV-A. The Benjamini–Hochberg false-discovery-rate correction was applied across this family of four (not across all $\binom{8}{2} = 28$ possible pairs, which would mix incomparable within-direction and cross-direction comparisons). Significance threshold $p_{\mathrm{BH}} < 0.05$.

##### S3.5 Platt scaling cross-validation

Platt scaling [11] was performed via 5-fold stratified cross-validation on the external test set. For each external validation run: the test cohort is partitioned into 5 outcome-stratified folds with `seed = 42`; for fold $k$, a fresh `LogisticRegression(penalty=None, solver="lbfgs", max_iter=1000)` is fit on the predicted-probability/outcome pairs of the other 4 folds and applied to fold $k$; the 5 held-out vectors are concatenated into a full-cohort recalibrated probability vector that is independent of any single calibrator fit. Calibration slope, intercept, observed-to-expected ratio, and Brier score are computed on the held-out vector. AUC is rank-invariant under the monotonic Platt transform, so AUC is reported pre-recalibration; calibration metrics are reported pre and post.

#### S4 Matched-subsampling sensitivity

This section gives the two-stratum maximum-$n$ subsampling pseudocode used to build matched-case-mix subsamples for the direction-asymmetry sensitivity analysis (main §2.4), worked numerical examples for each testable dimension, and the full three-framings-per-dimension results table referenced from main §2.4 (the headline range sentences therein cite this table for per-framing CIs).

##### S4.1 Algorithm

For a binary stratum vector and a target proportion of the positive stratum, the engine chooses between two subsampling options and returns indices for whichever option preserves more cases (*maximum-n*). Pseudocode is code-verified against the matched-subsampling implementation.

**Algorithm 2** Two-stratum maximum-$n$ matched subsampling.

**Require:** Stratum vector $s \in \{S^+, S^-\}^n$; positive level $S^+$; target proportion $p^* \in (0, 1)$
**Ensure:** Indices yielding $p^*$ proportion of $S^+$ at maximum $n$

$I^+ \leftarrow \{i : s_i = S^+\},\ \ I^- \leftarrow \{i : s_i \neq S^+\}$
$n^+ \leftarrow |I^+|,\ \ n^- \leftarrow |I^-|$
**Option A:** keep all $I^+$, subsample $I^-$ to $n_A^- = \text{round}(n^+(1 - p^*)/p^*)$
**if** $n_A^- \leq n^-$ **then** ▷ Option A feasible
    $\text{total}_A \leftarrow n^+ + n_A^-$
**else**
    $\text{total}_A \leftarrow -1$
**end if**
**Option B:** keep all $I^-$, subsample $I^+$ to $n_B^+ = \text{round}(n^- p^*/(1 - p^*))$
**if** $n_B^+ \leq n^+$ **then**
    $\text{total}_B \leftarrow n_B^+ + n^-$
**else**
    $\text{total}_B \leftarrow -1$
**end if**
**if** $\text{total}_A \geq \text{total}_B$ and $\text{total}_A > 0$ **then**
    Sample $n_A^-$ indices from $I^-$ without replacement; concatenate with all of $I^+$
**else**
    Sample $n_B^+$ indices from $I^+$ without replacement; concatenate with all of $I^-$
**end if**
**return** sorted index vector

The case-level paired bootstrap (§S3.1; 2,000 iterations, seed 42) is then run *on the matched subsamples* to compute the direction-asymmetry difference and its 95% percentile CI.

#### S4.2 Worked examples

**ASA stratum, framing A1 (match to INSPIRE's 90.4 / 9.6 case-mix).** INSPIRE is already at 90.4 / 9.6 ASA 1–2 / $\geq 3$, so no subsampling. MOVER has $n^+_{\text{ASA1-2}} = 21{,}028$ and $n^-_{\text{ASA}\geq 3} = 36{,}517$. Target $p^* = 0.904$. Option A keeps all 21,028 positives and subsamples negatives to round(21,028$\times$0.096/0.904) = 2,233 for total 23,261. Option B keeps all 36,517 negatives and would need round(36,517 $\times$ 0.904/0.096) = 343,990 positives, exceeding the available 21,028. Option A wins; matched MOVER size 23,261 (consistent with the matched-subsampling output).

**Emergency-case proportion, framing A1 (INSPIRE matched up to MOVER's 15.6% rate).** INSPIRE has 10,033 emergency cases and 117,380 non-emergency cases (7.9% emergency). Target $p^* = 0.156$. Option A keeps all 10,033 emergency and subsamples non-emergency to round(10,033 $\times$ 0.844/0.156) = 54,281 for total 64,314. Option B keeps all 117,380 non-emergency and would need round(117,380 $\times$ 0.156/0.844) $\approx$ 21,690 emergency, exceeding the available 10,033. Option A wins; matched INSPIRE size 64,314 (matches output). MOVER stays at 57,545 (already at 15.6%).

**Elixhauser comorbidity (Van Walraven), framing A1 (INSPIRE matched up to MOVER's 56.72% high-VW proportion at the pooled-median VW threshold of 1.00).** INSPIRE's high-VW count is below 56.72% of cohort. Option A subsamples INSPIRE non-high-VW cases until the high-VW proportion reaches 56.72%. Matched INSPIRE size is 107,247; MOVER stays at 57,545.

##### S4.3 Full results across three framings per dimension

Table S5 reports the matched direction-asymmetry across all three testable dimensions and three framings each, with their 95% percentile bootstrap CIs. The temporal-period dimension is recorded as infeasible (§S4.4). The headline range sentences in main §2.4 ("ASA: 128–203% of baseline", "Elixhauser: 113–126%", "emergency: 70–86%", "approximately 70% of baseline" for the emergency A1 framing) are derived as the Matched Δ in each row divided by the unmatched baseline of 8.53 pp.

Table S5: Matched direction-asymmetry across testable case-mix dimensions (main §2.4 sensitivity). Baseline (unmatched) direction asymmetry is 8.53 pp (95% CI 6.91–10.24; $p = 0.001$). All matched-framing $p$-values are 0.001 (the bootstrap Monte Carlo floor; §S3.2).

| **Dimension** | **Framing** | **Matched Δ (pp)** | **95% CI** | $n_{\mathbf{INS}}$ / $n_{\mathbf{MOV}}$ |
|---|---|---|---|---|
| ASA stratum | A1 (match INSPIRE 90.4/9.6) | 17.31 | 12.52–21.60 | 127,413 / 23,261 |
| | A2 (match MOVER 36.5/63.5) | 11.56 | 9.69–13.43 | 19,329 / 57,545 |
| | B (balanced 50/50) | 10.90 | 9.03–12.90 | 24,548 / 42,056 |
| Elixhauser comorbidity | A1 (match MOVER 56.72%) | 9.65 | 7.98–11.40 | 107,247 / 57,545 |
| | A2 (match INSPIRE 47.75%) | 10.76 | 8.95–12.52 | 127,413 / 47,658 |
| | B (balanced 50/50) | 9.68 | 7.93–11.58 | 121,670 / 49,806 |
| Emergency proportion | A1 (INSPIRE up to 15.6%) | 6.01 | 4.25–7.93 | 64,314 / 57,545 |
| | A2 (MOVER down to 7.9%) | 7.32 | 5.37–9.22 | 127,413 / 52,723 |
| | B (midpoint 11.75%) | 7.16 | 5.25–9.02 | 85,387 / 55,023 |
| Temporal period | — | Infeasible (§S4.4) | | |

##### S4.4 Why temporal matching is infeasible

Temporal-period matching was not attempted because INSPIRE distributes timestamps as privacy-preserving relative-time offsets (per Lim et al. 2024 [7]) and the release does not include a calendar-date crosswalk. Without an absolute reference date for any cohort member, the INSPIRE temporal axis cannot be aligned with MOVER's calendar dates (2015–2022); restricting both cohorts to a temporal-overlap window is therefore not runnable against the current INSPIRE release. This limitation is acknowledged in main §2.4 and §4.5 (Limitations).

#### S5 Calibration and inferential robustness

##### S5.1 Per-model calibration breakdown

Pre-recalibration calibration slopes ranged from 0.41–1.29 across the eight external validation runs and observed-to-expected (O:E) ratios were direction-asymmetric. INSPIRE-trained models on MOVER systematically under-predicted in-hospital mortality (O:E 0.02–0.10 across the four models). MOVER-trained models on INSPIRE separated by algorithm: logistic-regression variants under-predicted (O:E 0.10–0.19), while XGBoost variants over-predicted (O:E up to 2.92). Platt scaling, applied via the 5-fold stratified cross-validation procedure detailed in

§S3.5, restored calibration uniformly: post slopes 0.95–1.02, post O:E 0.99–1.01, mean Brier-score reduction of approximately 59.9% across the eight runs (Figures S1, S2; Table S6). Pre-recalibration reliability diagrams (Figure S3) make the direction-asymmetric miscalibration visible at decile resolution.

The calibration-analogue pattern mirrors the discrimination-asymmetry finding (§2.4): the same direction asymmetry that produces 2.6-fold greater external AUC degradation in MOVER-trained models also produces qualitatively different miscalibration by training direction. Discrimination and calibration both fail more severely when training on the higher-acuity cohort (MOVER) and deploying to the broader-spectrum cohort (INSPIRE), consistent with cross-population transfer where the absolute risk mapping learned from a concentrated case-mix does not generalize. AUC is rank-invariant under monotonic Platt scaling; the table reports AUC as a single column shared between the pre and post views.

Table S6: Per-model calibration metrics pre and post 5-fold cross-fit Platt scaling. Slope = 1.0, intercept = 0.0, and O:E = 1.0 are the well-calibrated targets. Pre-recal ranges and the direction-asymmetric pattern are visible across the eight pre rows. AUC is reported once per model on the pre row (rank-invariant under Platt; the corresponding post row carries a short-dash em-dash indicator).

| **Model** | **Stage** | **AUC** | **Slope** | **Intercept** | **O:E** | **Brier** |
|---|---|---|---|---|---|---|
| XGB-INS-A on MOVER | pre | 0.785 | 0.821 | −4.00 | 0.046 | 0.162 |
| | post | — | 0.999 | −0.00 | 1.003 | 0.014 |
| XGB-INS-B on MOVER | pre | 0.895 | 1.294 | −2.68 | 0.098 | 0.048 |
| | post | — | 1.004 | 0.01 | 0.998 | 0.013 |
| LR-INS-A on MOVER | pre | 0.796 | 0.713 | −4.69 | 0.032 | 0.282 |
| | post | — | 1.025 | 0.09 | 0.998 | 0.014 |
| LR-INS-B on MOVER | pre | 0.741 | 0.411 | −5.67 | 0.018 | 0.675 |
| | post | — | 0.949 | −0.22 | 0.996 | 0.014 |
| XGB-MOV-A on INSPIRE | pre | 0.756 | 0.507 | −1.51 | 1.355 | 0.011 |
| | post | — | 0.954 | −0.21 | 1.007 | 0.011 |
| XGB-MOV-B on INSPIRE | pre | 0.812 | 0.792 | 0.23 | 2.921 | 0.010 |
| | post | — | 0.955 | −0.21 | 0.999 | 0.010 |
| LR-MOV-A on INSPIRE | pre | 0.806 | 0.479 | −3.04 | 0.193 | 0.021 |
| | post | — | 0.995 | −0.02 | 1.003 | 0.010 |
| LR-MOV-B on INSPIRE | pre | 0.839 | 0.661 | −3.18 | 0.109 | 0.034 |
| | post | — | 1.012 | 0.04 | 0.995 | 0.010 |

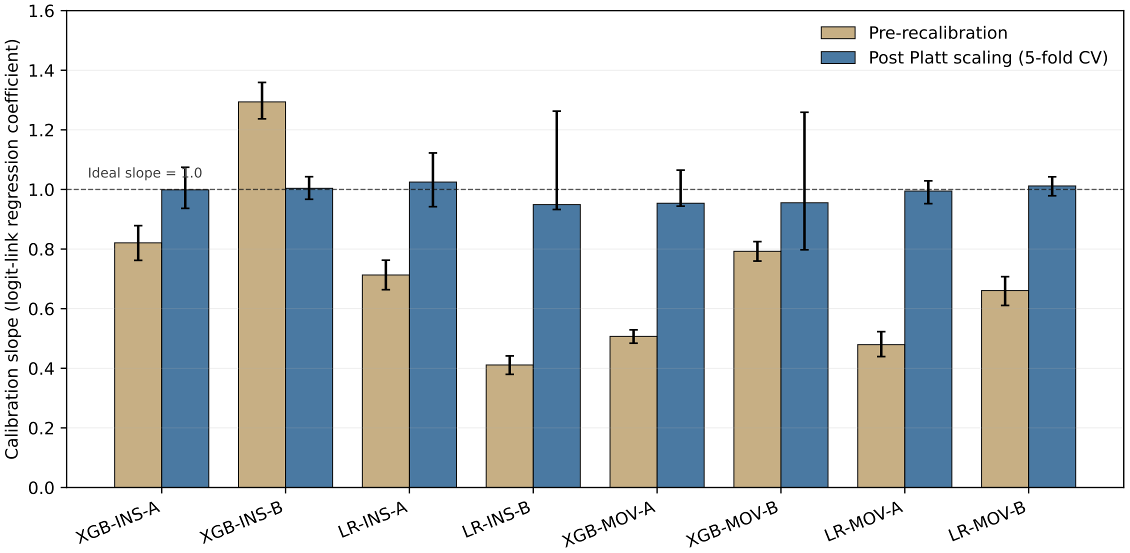

Figure S1: Per-model calibration slope before and after Platt scaling. Pre slopes (tan) span 0.41–1.29, with logistic-regression on the INSPIRE direction (LR-INS-B) at the lowest and XGBoost on the same direction (XGB-INS-B) at the highest. Post-recalibration slopes (blue) all include 1.0 within their 95% bootstrap CIs (0.95–1.02). Error bars are 2,000-resample case-level paired bootstrap CIs.

*Alt text: Paired-bar chart with eight model panels. For each model, two horizontal bars show calibration slope before Platt-scaling recalibration (tan, with whiskers at the 95% bootstrap CI) and after recalibration (blue, also with bootstrap CI). The pre-recalibration tan bars span a wide range from 0.41 (LR-INS-B, the worst) to 1.29 (XGB-INS-B, the best), all far from the ideal slope of 1.0. The post-recalibration blue bars cluster tightly between 0.95 and 1.02, with each 95% CI now including 1.0. The visualization makes immediately visible that Platt scaling restores calibration uniformly across the eight cross-population validation runs.*

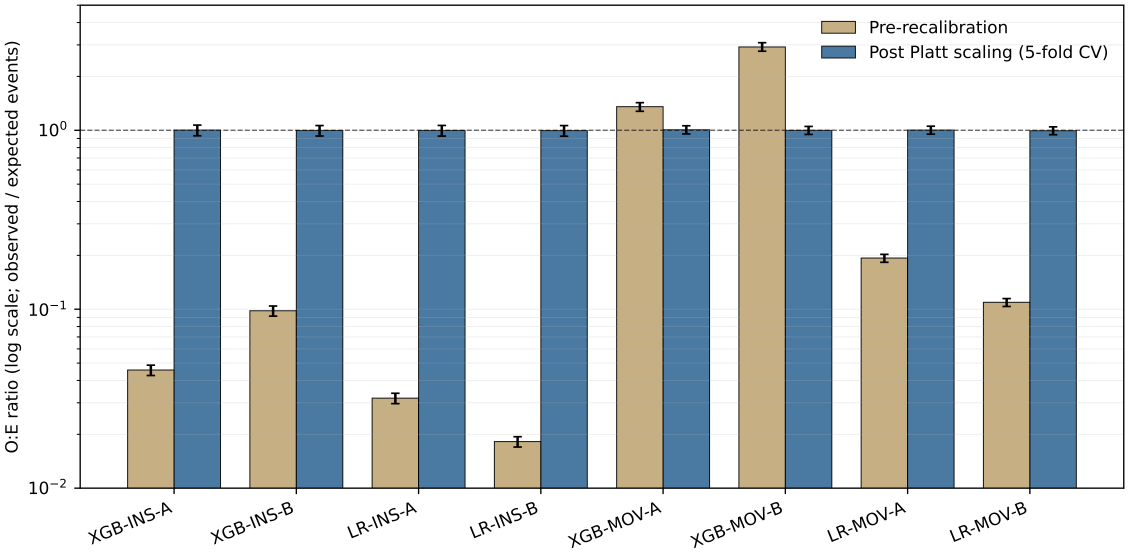

Figure S2: Per-model observed-to-expected (O:E) ratios before and after Platt scaling, on a logarithmic vertical axis (pre-recal range spans two orders of magnitude). The direction-asymmetric pre-recal pattern is visible: INSPIRE-trained on MOVER under-predicts (0.02–0.10); MOVER-trained on INSPIRE under-predicts (LR variants 0.10–0.19) or over-predicts (XGB variants up to 2.92). Post-recal: 0.99–1.01. Error bars are 95% bootstrap CIs.

*Alt text: Paired-bar chart with eight model panels on a logarithmic vertical axis (pre-recal range spans two orders of magnitude). For each model, two bars show observed-to-expected ratio (O:E) before Platt-scaling recalibration (tan) and after (blue), with 95% bootstrap CI whiskers. The pre-recalibration tan bars reveal a direction-asymmetric pattern: INSPIRE-trained models on MOVER under-predict mortality (O:E values clustered around 0.02 to 0.10, well below the ideal of 1.0); MOVER-trained models on INSPIRE either under-predict (LR variants, O:E around 0.10 to 0.19) or substantially over-predict (XGB variants, O:E up to 2.92). The post-recalibration blue bars cluster tightly around 1.0 (range 0.99–1.01) for every model. The visualization documents both the direction-asymmetric pre-recal mis-calibration and Platt scaling's uniform corrective effect.*

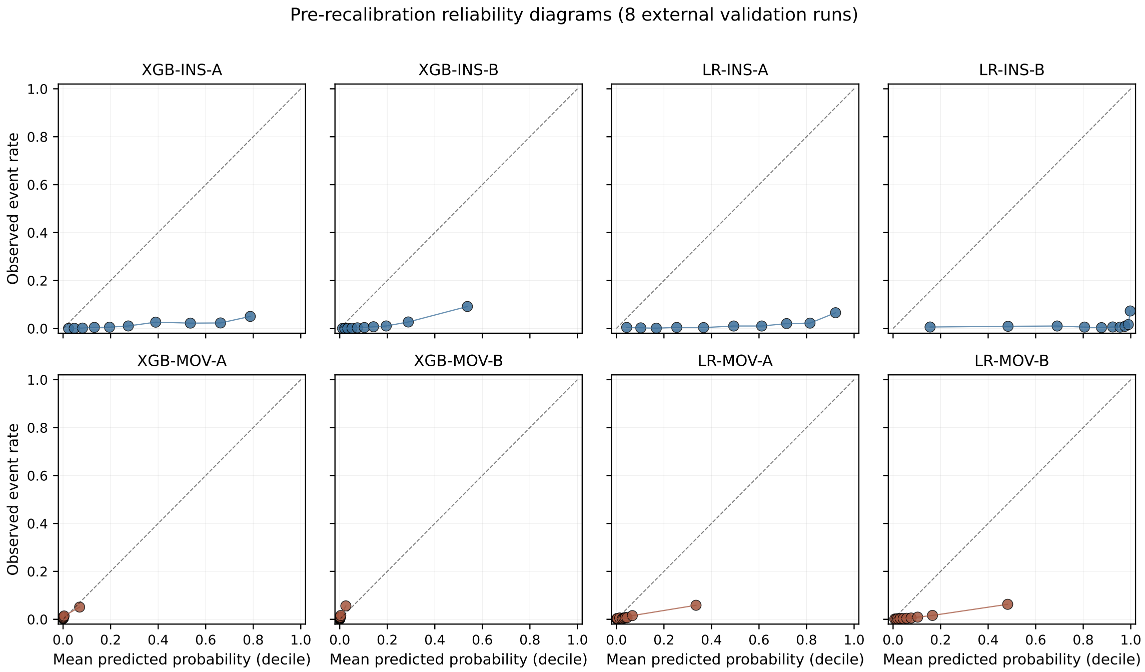

Figure S3: Pre-recalibration reliability diagrams for the eight external validation runs (deciles of predicted probability vs. observed event rate). Marker size scales with bin n. Top row: INSPIRE-trained models on MOVER (under-prediction visible as observed rates above the diagonal). Bottom row: MOVER-trained models on INSPIRE (XGB variants over-predict in the upper deciles; LR variants under-predict). The diagonal is the ideal calibration line.

*Alt text: Eight-panel reliability-diagram grid showing pre-Platt-scaling calibration for every external validation run. Within each panel, the horizontal axis is decile of predicted probability and the vertical axis is observed event rate; markers are placed per decile, sized by bin sample count, and the dashed diagonal is the ideal-calibration reference. Top row (INSPIRE-trained models on MOVER): markers consistently lie above the diagonal in the upper deciles, indicating under-prediction of mortality. Bottom row (MOVER-trained models on INSPIRE): the XGB variants show markers above the diagonal in the upper deciles (over-prediction); the LR variants show markers below the diagonal (under-prediction). The grid makes the direction- and algorithm-specific mis-calibration patterns visually inspectable per model.*

#### S5.2 Inferential robustness

The principal findings reported in main §2.3–§2.6 carry their own case-level paired bootstrap CIs (2,000 iterations, `seed = 42`; §S3.1); all bootstrap CIs and two-sided p-values are reproducible from the cached per-model predictions documented in main §3.6 without retraining. The permutation test on model-level degradation (4-vs-4 split) has minimum achievable $p = 1/70 \approx 0.014$ (§S3.3) and is floor-bounded irrespective of the underlying effect, which is why case-level inference is the primary framework. DeLong's asymptotic test (§S3.4) is reported as a within-direction sensitivity check across the four pairwise preop-vs-intraop comparisons; pairwise significance under Benjamini–Hochberg correction matches the case-level paired bootstrap conclusions in main §2.5. Bootstrap-iteration sensitivity (1,000 vs. 2,000 resamples) and seed sensitivity were spot-checked during development without producing meaningful point-estimate or CI shifts; comprehensive sweep tables are not distributed because the underlying conclusions are determined by case-count rather than resample-count, and the case counts (127,413 and 57,545) are large.

#### S5.3 Data-constrained analyses

Three analyses we would have run if the data permitted are reported as constraints rather than results.

**Race/ethnicity stratification.** The MOVER public release omits race/ethnicity at the source-data level: the `PATIENT_RACE_C` and `PATIENT_ETHNIC_C` fields are absent from the released MOVER EHR data tables (verified directly against the public release). INSPIRE has no equivalent variable. No race-stratified or ethnicity-stratified discrimination analysis is therefore possible from the data as released, in either direction, and no surrogate inference is attempted; the absence is documented as a data-availability constraint rather than a null result. This is a fairness limitation explicitly recommended-against by the clinical machine-learning fairness literature [3, 9] and is recorded as a primary limitation in main §4.5.

**Temporal period matching.** INSPIRE timestamps are privacy-preserving relative-time offsets without a calendar-date crosswalk; the 2015–2020 overlap window with MOVER cannot be enforced (§S4.4).

**Procedure-type matching.** INSPIRE's KCD-10 and MOVER's CPT/EPIC procedure code taxonomies do not align without a purpose-built crosswalk. Procedure-type subgroup matching is therefore not attempted.

#### S5.4 SHAP feature transferability

Figures S4 and S5 are the SHAP summary plots for the best XGBoost model in each transfer direction: XGB-INS-B externally validated on MOVER, and XGB-MOV-B externally validated on INSPIRE. SHAP-rank ordering is preserved across institutions with Spearman $\rho = 0.972$ and $\rho = 0.931$ across all 140 shared features (main §2.6); the top-ranked features are ASA, BMI, age, and heart-rate- derived intraoperative summaries in both directions (Table S7). The dissociation between rank-stable feature importance and direction-asymmetric calibration is the empirical anchor for the "relational structure transfers, absolute mappings do not" interpretation in main §4.3.

Table S7: Top-10 transferable features by SHAP rank for the best XGBoost model in each transfer direction. For each direction, internal rank (10-fold OOF) and external rank (held-out alternate cohort) are shown; rank-stability is high in both directions (Spearman $\rho$ across all 140 shared features: $\rho = 0.972$ for XGB-INS-B; $\rho = 0.931$ for XGB-MOV-B). ASA is rank-1 in both directions; demographic features (BMI, age) and HR-derived intraoperative summaries dominate the top 10.

| | **XGB-INS-B (INSPIRE → MOVER)** | | | **XGB-MOV-B (MOVER → INSPIRE)** | | |
|---|---|---|---|---|---|---|
| **Rank** | **Feature** | **Int.** | **Ext.** | **Feature** | **Int.** | **Ext.** |
| 1 | asa | 1 | 1 | asa | 1 | 1 |
| 2 | bmi | 2 | 2 | bmi | 2 | 3 |
| 3 | age | 3 | 4 | age | 3 | 4 |
| 4 | hr_mean | 4 | 5 | height_cm | 4 | 8 |
| 5 | sex | 5 | 8 | weight_kg | 5 | 5 |
| 6 | emergency | 6 | 6 | high_asa | 6 | 2 |
| 7 | hr_n | 7 | 3 | etco2_std | 7 | 6 |
| 8 | hr_min | 8 | 10 | high_asa_emergency | 8 | 9 |
| 9 | department | 9 | 12 | hr_first | 9 | 12 |
| 10 | dbp_art_mean | 10 | 13 | hr_mean | 10 | 7 |

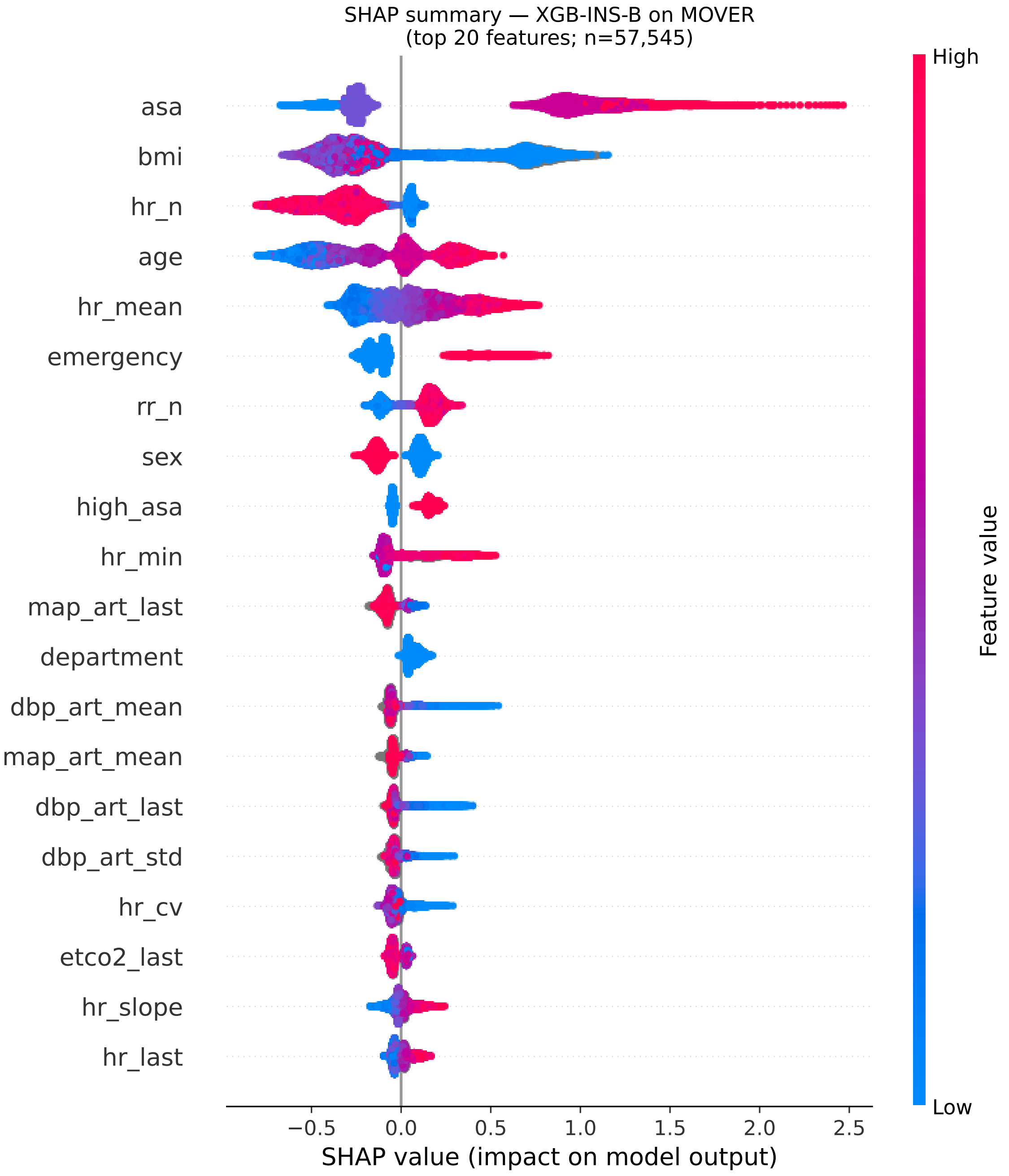

Figure S4: SHAP summary plot for XGB-INS-B externally validated on MOVER. Top-ranked features by mean absolute SHAP value: ASA, BMI, age, and heart-rate-derived intraoperative summaries. Internal-vs-external Spearman rank correlation across all 140 shared features: $\rho = 0.972$.

*Alt text: SHAP summary plot for the best XGBoost intraoperative model trained on INSPIRE and externally validated on MOVER. Vertical axis lists the top-ranked features by mean absolute SHAP value (ASA, BMI, age, then a sequence of heart-rate-derived intraoperative summaries). Horizontal axis is per-case SHAP value (impact on the model's log-odds prediction). Each row shows a swarm of points (one per MOVER patient), colored from blue (low feature value) through red (high feature value), so the plot reveals both feature importance ranking and per-feature directionality of effect. Internal-to-external Spearman rank correlation across all 140 shared features is reported in the figure caption; the rank ordering visible here is preserved relative to the model's INSPIRE training-set ranking.*

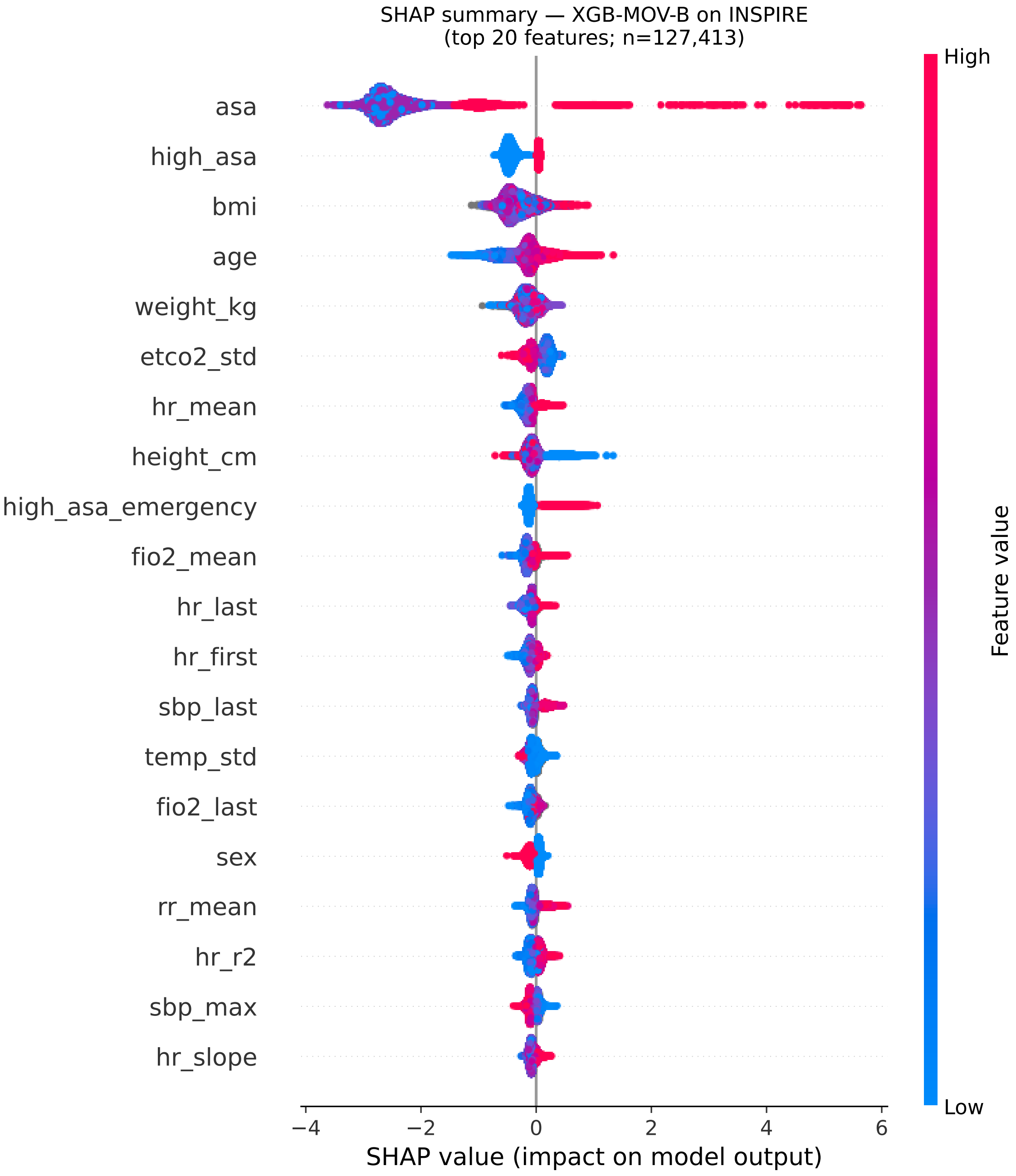

Figure S5: SHAP summary plot for XGB-MOV-B externally validated on INSPIRE. Top-ranked feature ordering matches XGB-INS-B's. Internal-vs-external Spearman rank correlation across all 140 shared features: $\rho = 0.931$.

*Alt text: SHAP summary plot for the best XGBoost intraoperative model trained on MOVER and externally validated on INSPIRE — the reverse direction of the previous figure. Vertical axis lists top-ranked features by mean absolute SHAP value, with the same dominant features (ASA, BMI, age, heart-rate-derived intraoperative summaries) as in the INSPIRE→MOVER direction. Horizontal axis is per-case SHAP value; each row shows a swarm of per-case impact dots colored by feature value (blue = low, red = high). The figure's central finding, when read alongside the previous figure, is that top-feature ordering is preserved across the bidirectional training/test swap.*

##### S5.5 Decision-curve clinical utility (per-direction detail)

Figure S6 reports per-model decision-curve net benefit across the 2–10% threshold range; the aggregated view is in main Figure 4. The per-model breakdown makes the LR-INS-B utility-loss above 2% threshold visible (consistent with its intraoperative-feature underperformance reported in main §2.5); the other seven models retain positive net benefit across the full 2–10% interval.

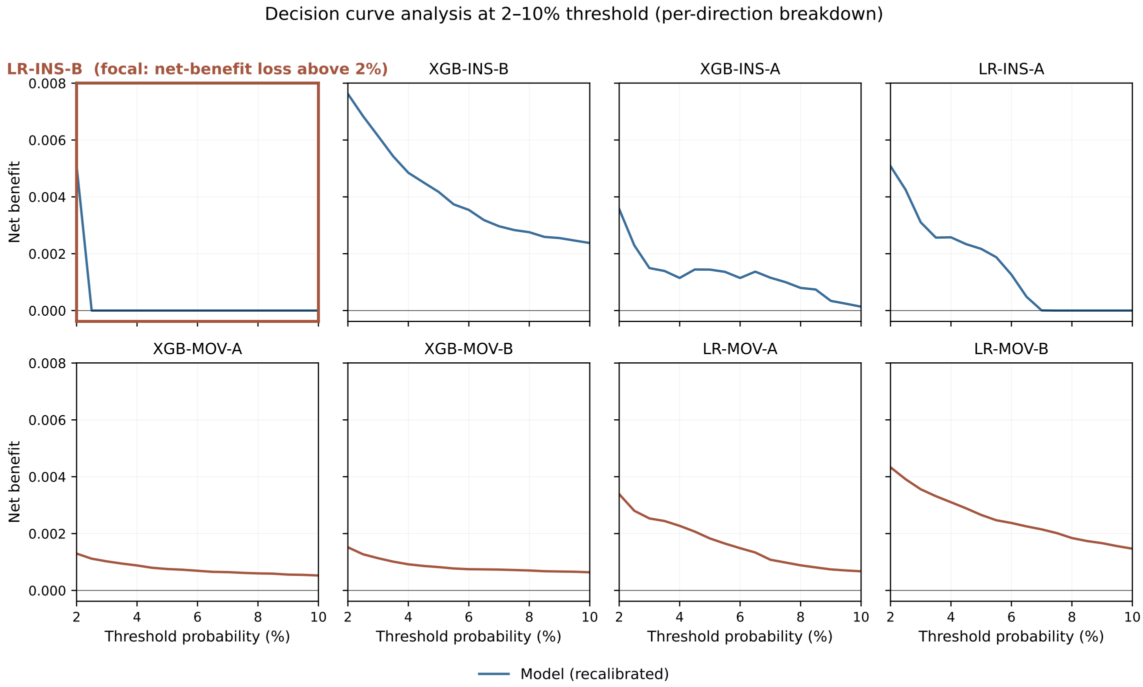

Figure S6: Per-model decision-curve net benefit at 2–10% threshold (post-Platt-scaling). Top row: INSPIRE-trained models on MOVER; bottom row: MOVER-trained models on INSPIRE. The LR-INS-B panel (top-left) is highlighted as the focal deployment-failure case (net-benefit loss above 2% threshold).

*Alt text: Eight-panel decision-curve grid showing post-Platt-scaling per-model net benefit across the clinically relevant 2%–10% decision threshold range. Top row: four INSPIRE-trained models on MOVER. Bottom row: four MOVER-trained models on INSPIRE. Within each panel, the model's net-benefit curve is plotted alongside the treat-all reference. Most panels show positive net benefit across the full threshold range. The top-left panel (LR-INS-B) is highlighted as the focal deployment-failure case: net benefit drops below the treat-all reference for thresholds above 2%, indicating that this specific model, despite recalibration, would harm clinical utility if deployed on MOVER for thresholds in the upper part of the clinically relevant range.*

#### S6 Aggregated statistics for verification

This section provides aggregated, claim-level reference tables that allow verification of every main-text quantitative claim without redistributed patient-level data. Each subsection's table is sourced from the canonical analysis outputs and is consistent with the values reported in main results; aggregated publication is consistent with both DUAs' minimum-cell-size requirements.

##### S6.1 Per-model per-stratum AUCs

Table S8 reports per-model overall and within-stratum external AUCs across two stratification axes (ASA stratum; sex). Cells marked [a] flag $n_{\text{events}} < 10$ in that stratum (single-stratum fallback applies in the paradox-gap calculation; main §2.3 + Table 2).

Table S8: Per-model external AUCs by stratum (overall, ASA 1–2, ASA $\geq$ 3, female, male). Cells marked [a] flag $n_{\text{events}} < 10$ (MOVER's ASA 1–2 stratum has 9 deaths).

| **Model** | **Overall** | **ASA 1–2** | **ASA $\geq 3$** | **Female** | **Male** |
|---|---|---|---|---|---|
| XGB-INS-A on MOVER | 0.785 | 0.621[a] | 0.682 | 0.783 | 0.775 |
| XGB-INS-B on MOVER | 0.895 | 0.724[a] | 0.845 | 0.893 | 0.890 |
| LR-INS-A on MOVER | 0.796 | 0.645[a] | 0.715 | 0.783 | 0.791 |
| LR-INS-B on MOVER | 0.741 | 0.858[a] | 0.687 | 0.737 | 0.733 |
| XGB-MOV-A on INSPIRE | 0.756 | 0.597 | 0.584 | 0.748 | 0.755 |
| XGB-MOV-B on INSPIRE | 0.812 | 0.705 | 0.741 | 0.830 | 0.796 |
| LR-MOV-A on INSPIRE | 0.806 | 0.685 | 0.683 | 0.809 | 0.784 |
| LR-MOV-B on INSPIRE | 0.839 | 0.746 | 0.771 | 0.848 | 0.819 |

[a]MOVER ASA 1–2 stratum has $n_{\text{events}} = 9$; AUC reported for completeness, not used in paradox gap.

##### S6.2 Per-model paradox gap summary

Table S9 is the paradox gap reference cited from main §4.4. It is the same data underlying main Table 2 (values match exactly); the difference is column structure: this table reports the gap and its CI in compact form for verification.

Table S9: Per-model Simpson's paradox gap summary.

| **Model** | **Trained on** | **Overall AUC** | **Within-stratum AUC** | **Gap (pp)** | **95% CI** |
|---|---|---|---|---|---|
| XGB-INS-A | INSPIRE | 0.785 | 0.682 | 10.3 | 9.6–10.9 |
| XGB-INS-B | INSPIRE | 0.895 | 0.845 | 5.0 | 4.4–5.6 |
| LR-INS-A | INSPIRE | 0.796 | 0.715 | 8.1 | 7.5–8.8 |
| LR-INS-B | INSPIRE | 0.741 | 0.687 | 5.4 | 4.9–5.8 |
| XGB-MOV-A | MOVER | 0.756 | 0.590 | 16.5 | 15.1–18.0 |
| XGB-MOV-B | MOVER | 0.812 | 0.723 | 8.9 | 7.9–10.0 |
| LR-MOV-A | MOVER | 0.806 | 0.684 | 12.2 | 10.7–13.6 |
| LR-MOV-B | MOVER | 0.839 | 0.759 | 8.0 | 6.8–9.2 |

##### S6.3 Per-model calibration metrics

The full per-model pre/post Platt calibration metrics (slope, intercept, O:E, Brier) are in Table S6 (§S5.1). That table is the verification-form aggregation backing the 0.41–1.29, 0.95–1.02, 0.99–1.01, and 59.9% summary statistics in main §2.6.

##### S6.4 Matched-sensitivity full values

Table S10 expands Supplementary Table S5 (§S4.3) with per-direction degradation columns (the matched mean degradation in INSPIRE-trained vs MOVER-trained groups, separately) that the asymmetry-only summary in S5 does not show.

Table S10: Matched direction-asymmetry full values per dimension and framing. Per-direction degradation columns are mean external AUC degradation across the four models in each training direction (relative degradation). The asymmetry column is MOVER-trained mean − INSPIRE-trained mean.

| Dimension | Framing | INS-deg | MOV-deg | Asym (pp) | 95% CI |
|---|---|---|---|---|---|
| Baseline | unmatched | 0.054 | 0.139 | 8.53 | 6.91–10.24 |
| ASA | A1 (INS 90.4/9.6) | −0.034 | 0.139 | 17.31 | 12.52–21.60 |
| ASA | A2 (MOV 36.5/63.5) | 0.054 | 0.170 | 11.56 | 9.69–13.43 |
| ASA | B (50/50) | 0.023 | 0.132 | 10.90 | 9.03–12.90 |
| Elixhauser | A1 (MOV 56.7%) | 0.054 | 0.150 | 9.65 | 7.98–11.40 |
| Elixhauser | A2 (INS 47.8%) | 0.054 | 0.162 | 10.76 | 8.95–12.52 |
| Elixhauser | B (50/50) | 0.054 | 0.151 | 9.68 | 7.93–11.58 |
| Emergency | A1 (INS up to 15.6%) | 0.054 | 0.114 | 6.01 | 4.25–7.93 |
| Emergency | A2 (MOV down to 7.9%) | 0.054 | 0.127 | 7.32 | 5.37–9.22 |
| Emergency | B (midpoint 11.75%) | 0.054 | 0.126 | 7.16 | 5.25–9.02 |
| Temporal | — | Infeasible (§S4.4) | | | |

#### S6.5 Sex-stratified AUCs

Per-model sex-stratified external AUCs with 95% bootstrap CIs and $|F - M|$ differential. The 5-pp threshold pre-specified for fairness flags (main §2.7) was not exceeded by any model.

Table S11: Per-model sex-stratified external AUCs. $|F - M|$ pp is the absolute female-male AUC differential in percentage points.

| Model | Female AUC (95% CI) | $n_F$ events | Male AUC (95% CI) | $n_M$ events | $\|F-M\|$ pp |
|---|---|---|---|---|---|
| XGB-INS-A on MOVER | 0.783 (0.761–0.803) | 264 | 0.775 (0.757–0.792) | 559 | 0.8 |
| XGB-INS-B on MOVER | 0.893 (0.875–0.908) | 264 | 0.890 (0.878–0.901) | 559 | 0.3 |
| LR-INS-A on MOVER | 0.783 (0.754–0.812) | 264 | 0.791 (0.772–0.810) | 559 | 0.7 |
| LR-INS-B on MOVER | 0.737 (0.697–0.776) | 264 | 0.733 (0.707–0.757) | 559 | 0.4 |
| XGB-MOV-A on INSPIRE | 0.748 (0.723–0.771) | 520 | 0.755 (0.736–0.773) | 867 | 0.7 |
| XGB-MOV-B on INSPIRE | 0.830 (0.811–0.849) | 520 | 0.796 (0.780–0.811) | 867 | 3.4 |
| LR-MOV-A on INSPIRE | 0.809 (0.788–0.830) | 520 | 0.784 (0.767–0.800) | 867 | 2.6 |
| LR-MOV-B on INSPIRE | 0.848 (0.828–0.865) | 520 | 0.819 (0.804–0.833) | 867 | 2.8 |

#### S6.6 Bootstrap iteration-count sensitivity

The headline direction-asymmetry case-level paired bootstrap was re-run at $B = 500$ and $B = 1{,}000$ in addition to the canonical $B = 2{,}000$ (Table S12). Point estimates and CI bounds are stable across resample counts; CI width narrows as $B$ increases but the inferential conclusion (asymmetry $> 0$ at the Monte Carlo floor $p = 0.001$, §S3.2) does not depend on the resample count at the case-level $n$ used here.

Table S12: Bootstrap iteration-count sensitivity for the headline direction- asymmetry claim, computed at $B = 500$, $B = 1{,}000$, and $B = 2{,}000$ from the cached per-model predictions using the paired-bootstrap engine described in §S3.1.

| $B$ | **Point estimate (pp)** | **95% CI lower (pp)** | **95% CI upper (pp)** | **CI width (pp)** |
|---|---|---|---|---|
| 500 | 8.52 | 6.84 | 10.11 | 3.27 |
| 1,000 | 8.53 | 6.84 | 10.19 | 3.35 |
| 2,000 | 8.53 | 6.85 | 10.25 | 3.40 |

### S7 Healthcare-system mechanism evidence

This section provides the supporting evidence for the healthcare-system mechanism interpretation introduced in main §4.3. The data anchor (a 50.6-pp split-difference between INSPIRE's spectrum-distributed and MOVER's high-acuity-concentrated mortality patterns at matched ASA stratum) is established in main §4.3; here we report the 2×2 stratified mortality table that backs it (§S7.1) and expand the cross-institutional end-of-life-practice literature that grounds the healthcare-system-specific feature–outcome-mapping interpretation (§S7.2). Cross-references to the matched-case-mix analysis are in §S7.3.

#### S7.1 Stratified mortality distribution

The full distribution of in-hospital deaths by ASA stratum across the two cohorts is in Table S13. INSPIRE deaths spread across the acuity spectrum (47.2% in ASA 1–2 vs 52.8% in ASA $\geq$ 3), whereas MOVER deaths concentrate almost entirely in high-acuity patients (1.1% in ASA 1–2 vs 98.9% in ASA $\geq$ 3). The corresponding within-stratum mortality rates also diverge markedly: INSPIRE ASA 1–2 rate 0.57% (654 deaths over 115,139 (90.4%) surgeries) vs MOVER ASA 1–2 rate 0.04% (9 deaths over 21,028 (36.5%)); INSPIRE ASA $\geq$ 3 rate 5.97% (733 deaths over 12,274 (9.6%)) vs MOVER ASA $\geq$ 3 rate 2.23% (814 deaths over 36,517 (63.5%)). The mortality-share split between the two strata is therefore not a small perturbation; it is structural.

Table S13: Stratified in-hospital mortality distribution by cohort and ASA stratum. Cells report deaths $n$, share-of-deaths within cohort, and within-stratum mortality rate.

| **Cohort** | **ASA 1–2 deaths** | **ASA $\geq$ 3 deaths** |
|---|---|---|
| INSPIRE | 654 (47.2%; 0.57%) | 733 (52.8%; 5.97%) |
| MOVER | 9 (1.1%; 0.04%) | 814 (98.9%; 2.23%) |

#### S7.2 Cross-institutional end-of-life practice patterns

End-of-life decision-making practices in intensive care units differ systematically between East Asian and Western settings, and these differences may shape the joint distribution of physiological trajectories and observed in-hospital mortality outcomes. Phua and colleagues [10] surveyed end-of-life practices across 16 Asian countries and territories (including South Korea), reporting that the proportion of deaths preceded by withholding or withdrawing life-sustaining treatments was substantially lower than that documented in contemporaneous Western studies, and that withdrawing (as distinct from withholding) was particularly uncommon in many Asian sites. Within South Korea specifically, Park and colleagues [8] described sharp temporal shifts in withdrawing/withholding practices coincident with the introduction of the Act on Hospice and Palliative Care (implemented 2018), and documented that, even after the legal change, attending-physician discretion and family-centric decision frameworks remained associated with substantial inter-institutional variability in EOL care patterns. The 50.6-pp split-difference between INSPIRE's spectrum-distributed and MOVER's high-acuity-concentrated mortality patterns (main §4.3; §S7.1) is consistent with this class of practice variability, though our data alone cannot identify any specific mechanism.

##### S7.3 Implication for the residual asymmetry

The matched-case-mix sensitivity analysis (§S4) established that approximately 70% of the unmatched direction-asymmetry baseline survives matching on three testable case-mix dimensions (ASA, Elixhauser comorbidity, emergency-case proportion). This residual asymmetry is consistent with healthcare-system-specific feature–outcome relationships that case-mix matching does not equalize — both because some sources of asymmetry (temporal period; §S4.4) cannot be matched, and because some sources operate within strata in ways inter-rater-variable ASA classification [13] does not capture. The cross- institutional EOL-practice patterns reviewed in §S7.2 are one plausible mechanism class among several; our cohort data cannot directly distinguish among candidate mechanisms. The interpretation in main §4.3 is therefore framed as "consistent with" rather than "established by" the available data.

#### S8 TRIPOD+AI reporting checklist

This section maps the manuscript against the TRIPOD+AI reporting checklist (Collins et al. 2024) [4]. Items 1–27 cover the full development–evaluation lifecycle; this study is an evaluation study (cross-cohort external validation of pre-trained models from INSPIRE / MOVER public data releases). For items that primarily concern model development, the relevant content is reported in Methods §3.2 + Supplementary §S2 (which document the development pipeline our analyses re-use).

Table S14: TRIPOD+AI reporting checklist (Collins et al. 2024) mapped to manuscript sections. "Main §X" refers to canonical Methods/Results/ Discussion sections; "Supp §Y" refers to numbered subsections in this supplementary file.

| Item | Topic | Manuscript reference |
|---|---|---|
| **Title and Abstract** | | |
| 1 | Identify the study as developing/evaluating a prediction model | Title; Abstract Objective |
| 2 | Structured abstract (Objective, Methods, Results, Conclusion) | Abstract |
| **Introduction** | | |
| 3a | Healthcare context and rationale | Main §1 (Background) |
| 3b | Target population, intended purpose, intended users | Main §1; §4.4 (Clinical implications) |
| 3c | Known health inequalities between sociodemographic groups | Main §4.5 (Limitations); Supp §S5.3 (Race/ethnicity unavailability) |

*(continued on next page)*

*(continued from previous page)*

| Item | Topic | Manuscript reference |
|---|---|---|
| 4 | Study objectives (development vs evaluation) | Main §1 closing paragraph; this is an evaluation study |
| **Methods** | | |
| 5a | Data sources, separately for dev / eval, with representativeness | Main §3.1; Supp §S1.1, §S1.2 |
| 5b | Dates of data collection (accrual + follow-up) | Main §3.1 (INSPIRE 2011–2020; MOVER 2015–2022) |
| 6a | Study setting, number/location of centres | Main §3.1 (single-centre each cohort) |
| 6b | Eligibility criteria for participants | Main §3.1; Supp §S1.1, §S1.2; main Figure 1 |
| 6c | Treatments received and how handled | N/A (cross-cohort EHR-based study; no treatment intervention) |
| 7 | Data pre-processing and quality checks across groups | Main §3.1; Supp §S2.1 (feature inventory + completeness rates) |
| 8a | Outcome definition, time horizon, assessment method | Main §3.1 (in-hospital mortality) |
| 8b | Outcome assessor qualifications (if subjective) | N/A (administrative outcome ascertainment) |
| 8c | Outcome assessment blinding | N/A |
| 9a | Predictor pre-selection before model building | Main §3.2; Supp §S2.1 (8 preop, 132 intraop) |
| 9b | Predictor definitions, measurement timing, blinding | Main §3.2 (60-min post-induction window); Supp §S2.1 |
| 9c | Predictor assessor qualifications | N/A (sensor-derived data) |
| 10 | Sample size determination and justification | Main §3.2 (EPV ratios stated inline) |
| 11 | Missing data handling | Main §3.2; Supp §S2.1 (intraop coverage 99.9% INSPIRE / 92.1% MOVER); §S2.2 (mean imputation for LR) |
| 12a | Data partitioning for analyses | Main §3.2 (10×5 nested CV); Supp §S2.2 |
| 12b | Predictor handling (transformation, scaling) | Supp §S2.2 (LR standardisation; XGBoost native handling) |
| 12c | Model type, building steps, hyperparameter tuning | Main §3.2; Supp §S2.2 (full search space + median tuned) |
| 12d | Cluster heterogeneity handling | N/A (no clustered structure beyond cohort) |
| 12e | Performance evaluation measures | Main §3.3, §3.5; Supp §S3, §S5 |
| 12f | Model updating during evaluation | N/A (frozen models; only Platt recalibration; Supp §S3.5) |
| 12g | Prediction calculation for evaluation | Main §3.5; Supp §S3.5 |
| 13 | Class imbalance methods + recalibration | Main §3.2 + §3.7; Supp §S2.3, §S3.5 |

*(continued on next page)*

*(continued from previous page)*

| Item | Topic | Manuscript reference |
|---|---|---|
| 14 | Model fairness approaches | Main §2.7 (sex stratification); Supp §S5.3, §S6.5 |
| 15 | Model output type and threshold rationale | Main §3.5; main §2.7 (DCA at 2–10%) |
| 16 | Differences between development and evaluation data | Main §2.1, §2.4; Table 1 |
| 17 | Ethics committee and consent | Main §3.1 (public credentialed-access datasets; separate IRB approvals at source institutions) |
| **Open Science** | | |
| 18a | Funding source and funder role | Main Acknowledgments / Funding [forthcoming at submission] |
| 18b | Conflicts of interest | Main Disclosures [forthcoming at submission] |
| 18c | Study protocol availability | Main §3.6 (no pre-registered protocol; canonical analysis pipeline + scripts under MIT) |
| 18d | Study registration | Not registered (post-hoc external-validation study of public datasets) |
| 18e | Data availability | Main §3.6 (DUA-Posture-2 commitment; Supp §S6 aggregated tables) |
| 18f | Analytical code availability | Main §3.6; Zenodo deposit (DOI assigned at submission) |
| **Patient and Public Involvement** | | |
| 19 | PPI in design / conduct / reporting | N/A (secondary use of pre-existing public datasets) |
| **Results** | | |
| 20a | Participant flow, outcome numbers, follow-up | Main §2.1; main Figure 1; Table 1 |
| 20b | Characteristics overall and by data source | Table 1 |
| 20c | Comparison of development vs evaluation data distribution | Main §2.1; Supp §S6.1 |
| 21 | Participant and event numbers in each analysis | Tables 2 + 3 + S8 |
| 22 | Full prediction model details (enabling third-party use) | Main §3.6; Supp §S2 + §S6; Zenodo |
| 23a | Performance with CIs, including key subgroups | Main §2.3–§2.7; Supp §S5, §S6.1, §S6.5 |
| 23b | Heterogeneity across clusters | Main §2.4 (matched-case-mix); Supp §S4 |
| 24 | Model updating results (if any) | Main §2.6 (Platt recalibration); Supp §S5.1, §S6.3, Table S6 |
| **Discussion** | | |
| 25 | Overall interpretation, fairness, prior studies | Main §4.1, §4.2, §4.3 |
| 26 | Limitations, bias, uncertainty, generalisability | Main §4.5; Supp §S5.3 |
| 27a | Handling of poor-quality input data at deployment | Main §4.4 (recalibration on representative deployment sample) |

*(continued on next page)*

*(continued from previous page)*

| Item | Topic | Manuscript reference |
|---|---|---|
| 27b | User interaction with input data | Main §4.4 (TRIPOD+AI-aligned reporting commitments) |
| 27c | Next steps for research | Main §4.4, §4.5 |
